# Factors associated with severity of pulmonary tuberculosis in people living with and without HIV: a systematic review

**DOI:** 10.1101/2025.07.06.25330980

**Authors:** Robert Akpata, Anani Badjé, Jean-Baptiste Ntakpé, Cossi Angelo Attinsounon, Nathalie de Castro, Didier Laureillard, Marcel Zannou, Olivier Marcy

## Abstract

**Introduction:** Tuberculosis is a leading infectious cause of death and morbidity in people with and without HIV. Better identification of severe forms could help target early interventions to reduce mortality and severe morbidity. Existing severity scores do not take into account important disease characteristics such as bacillary load, extent of lung involvement, and disease dissemination, and most of them were not developed specifically for people living with HIV. We sought to identify factors associated with tuberculosis severity described in the literature in order to propose a consensual multifactorial tuberculosis severity score for people living with HIV.

**Methods:** The review was conducted using the Preferred Reporting Items for Systematic Reviews and Meta-Analyses (PRISMA) approach. We searched for baseline data associated with tuberculosis severity defined as mortality, severe morbidity, treatment failure or sequelae, in participants aged ≥15 years.

**Results:** In 117 articles selected, the most commonly reported severity factors were host-related or mixed (older age, anaemia, immunosuppression, comorbidities, low body mass index, fever, tachycardia, dyspnoea, low blood pressure, or social factors). There were also factors related to bacillary load (high smear positivity level or culture positivity), extent of lung involvement (cavitation, bilateral location, or infiltration to more than 2 lobes) and disease dissemination (LAM+, extra pulmonary involvement, miliary or disseminated tuberculosis).

**Conclusion:** This review identified many factors associated with tuberculosis severity, host-related or related to both the host and tuberculosis. It also brought evidence that TB severity is related as well to key disease characteristics such as bacillary load, extent of lung involvement and disease dissemination. It will be used to develop a specific tuberculosis severity score for people living with HIV based on the Rand Appropriateness Method.

The systematic review protocol was registered in the Prospective Register of Systematic Reviews (PROSPERO, CRD42022323983).

Clinical trial number: not applicable.

## Introduction

Tuberculosis (TB) is the leading cause of death from a single infectious agent with 1.25 million deaths estimated in 2023 (1). Overall, WHO estimates that 10.8 million people fell ill with TB in 2023, experiencing in an unknown proportion significant morbidity (hospitalization, prolonged hospitalization, complications, admission to intensive care, treatment failure) (1). In people living with Human Immunodeficiency Virus (PLWH), TB is the leading cause of death, accounting for 161 000 deaths in 2023, and the leading opportunistic infection, with 662 000 estimated TB disease occurring in PLWH the same year (1). In addition, a large proportion of PLWH experience significant associated morbidity requiring hospitalization or prolonged hospitalization, paradoxical clinical deterioration due to Immune Reconstitution Inflammatory Syndrome (IRIS), delayed cure, treatment failure or relapse (2–7). Both people with and without HIV also face a risk of post-TB lung disease with approximately one-third to one-half of TB survivors affected, HIV negative and PLWH alike (8–14). As such, both in PLWH and people without HIV, TB is a severe disease, not only due to its lethality but also to risk of significant morbidity, as well as to the risk of long-term sequelae, typically post-TB lung disease (14–17).

Identifying individuals with more severe TB, i.e., higher risks of death, severe morbidity including treatment failure, and long term sequelae could lead to differentiated treatment options for patients based on disease severity (18,19). Individuals admitted with HIV-associated tuberculosis are frequently acutely ill, with high inpatient deaths rates occurring early during hospitalization, at a median of 4–5 days after admission (20–22), despite appropriate antituberculosis treatment (20). This suggests also the urgent need for acute management strategies to increase survival in PLWH with TB, including for instance higher doses of rifampicin or other intensified treatment regimens (23), which can be achieved only through a better understanding of the main contributors to severe morbidity and mortality.

Several scores have been previously proposed to assess TB severity. Based on clinical, biological, or radiological criteria, most of them apply to the general population, and very few specifically to PLWH. The Bandim TB score predicts 8 months mortality in TB patients from low-resource settings based on 5 symptoms (cough, hemoptysis, dyspnea, chest pain, night sweating) and 6 signs (anemia, tachycardia, abnormal lung auscultation, fever, low body-mass index (BMI) and low mid-upper arm circumference (MUAC)) (24). The simplified TB score II predicts 6 months mortality based on cough, dyspnea, chest pain, anemia, low BMI and low MUAC (25). The Timika score grades chest X-ray (CXR) severity and predicts treatment outcomes in patients with smear-positive pulmonary TB based on the percentage of lung affected and presence of cavitation (26). The CABI score predicts mortality during TB treatment based on clinical form of TB, age, adjusted BMI, and HIV infection (27). The Gupta-Wright score predicts 2-months mortality in PLWH based on age, sex, ongoing antiretroviral treatment (ART), anemia, inability to walk unaided and lipoarabinomannan (LAM) test result (28). The Nguyen and Graviss score, developed specifically for US-based TB patients predicts mortality based on age, US born status, homelessness, residency in long-term care facility, chronic kidney failure, meningeal TB (TBM), miliary TB, TB-specific abnormality on CXR and HIV status (29).

While the association of bacillary load, extent of lung involvement on CXR, and disease dissemination with endpoints characterizing disease severity is well established (3,30–33), these key TB disease characteristics are not taken into account in existing scores, at the exception of the Timika score, that includes CXR features, the Gupta-Wright score that includes urine LAM test as a marker of disease dissemination, and the Nguyen and Graviss score that may not apply in high TB burden resource-limited settings. None of the existing TB severity scores included these three characteristics, and thus might be sub-optimal, and might not be applicable to PLWH.

Aiming to develop a better tuberculosis severity score for PLWH, we sought to identify factors associated with severity of pulmonary TB described in the literature, notably those related to bacillary load, extent of lung involvement, and disease dissemination, and to assess differences in factors reported in PLWH and people without HIV.

## Methods

We conducted a systematic review as the first step of a consensus process aiming to develop a multifactorial TB severity score for PLWH including clinical criteria, bacillary load, extent of lung involvement on CXR and the disease dissemination, using the *RAND Appropriateness Method* (RAM), which combines two knowledge synthesis techniques: the literature review and the Delphi process (34). We followed the Preferred Reporting Items for Systematic Reviews and Meta-Analyses (PRISMA) checklist (35).

### Eligibility criteria

We selected publications reporting on severity scores, scales, risk factors associated with TB disease severity defined as either death, severe morbidity (hospitalization, prolonged hospitalization, complications, admission to intensive care, treatment failure), unsuccessful treatment (treatment default, treatment failure, death) or sequelae (long term respiratory impairment) as main outcomes. The variables of interest were baseline clinical, radiological, biological, and microbiological data associated with the outcomes and part of the standard of care. Participants were patients at least 15 years old with TB (pulmonary associated or not with extra pulmonary TB, new or relapse, drug-sensitive (DS) or drug-resistant (DR) TB), from all regions and countries, with HIV or not. There was no restriction based on study design, study setting, or duration of follow-up.

We included studies published in English, French, German or Spanish from 01 January 2010 to 31 October 2021. We excluded studies that reported only on extra pulmonary TB as well as studies with no numerical data regarding strength of association and those with univariate analysis only. Case reports, case series, scientific correspondences, and conference abstracts were also excluded.

### Information sources

We searched PubMed, Embase and Cochrane. To ensure completeness, we also checked the reference lists of included papers for additional relevant references.

### Search strategies

Specific search equations for each electronic database were developed using the keywords described in the supplementary appendix 1. The search equations in the different databases were developed by one author (RA) and validated by a librarian of the University of Bordeaux specialized in bibliographic research in health sciences (supplementary appendix 2.). The results of the bibliographic search were managed with Zotero software.

### Studies selection and data collection

The screening of articles by titles was completed by the first author (RA). Then the first author and a group of four senior researchers (AB, JBN, ACA and OM) independently reviewed abstracts for eligibility criteria. Finally, data from the selected publications were extracted by the first author and then reviewed and validated by a group of three senior researchers (AB, JBN and OM). At the end of each step, the results were compared, and disagreements resolved during meetings. Data were extracted into a Microsoft Excel Office 365 spreadsheet (Microsoft, Redmond, Washington, USA). The data extraction spreadsheet was pilot tested and refined before extraction (supplementary appendix 3.).

### Data items

The variables extracted from selected articles were: general information (DOI, first author, year of publication, language), objective, methods (country and context, population, study design), study results (population size, patients characteristics (age, sex, TB type, HIV status), duration of follow-up, outcomes, tuberculosis severity factors, summary measures of association (adjusted odds or hazard ratios, or relative risk), risk of bias (not serious, serious, very serious) and quality of evidence (supplementary appendix 3.).

### Risk of bias in individual studies

The overall risk of bias for each severity factor was determined using the *Grading of Recommendations Assessment, Development and Evaluation* (GRADE) classification (not serious, serious, very serious) by assessing if there were : i) failure to develop and apply appropriate eligibility criteria (under- or overmatching in case-control studies, selection of exposed and unexposed in cohort studies from different populations); ii) flawed measurement of both exposure and outcome (differences in measurement of exposure, differential surveillance for outcome in exposed and unexposed in cohort studies); iii) failure to adequately control confounding (failure of accurate measurement of all known prognostic factors, failure to match for prognostic factors and/or lack of adjustment in statistical analysis); iv) incomplete or inadequately short follow-up (36).

### Severity factors categorization

In an effort to highlight the role played by TB itself in the disease severity, we split the severity factors into three categories based on a questionnaire submitted to a panel of 17 TB experts: i) intrinsic factors (related to TB only; ii) extrinsic factors (related to the host only; iii) mixed factors (host and TB related). We also categorized variables as related to the 3 key diseases characteristics: i) bacillary load (smear positivity or high positivity level, culture positivity and time to culture positivity itself, Xpert positivity and level of MTB detection), ii) extent of lung involvement (cavitation, bilateral location, infiltration to more than 2 lobes, …/…) or iii) disease dissemination (LAM positivity, extra pulmonary involvement, miliary or disseminated TB).

### Summary measures and quality of evidence

We collected adjusted hazard ratios (aHR), adjusted odds ratios (aOR) and adjusted relative risks (aRR). Effect estimates and their confidence intervals were presented using forest plots. Severy factors associated with each outcome were summarized in tables. The quality of evidence of individual severity factors was assessed using the GRADE system (37). All severity factors started as low-quality evidence and were graded upward if: i) the magnitude of the association was large (adjusted relative risk (aRR), adjusted hazard ratio (aHR) or adjusted odds ratio (aOR) >2 or <0.5) or very large (aRR, aHR ora OR >5 or <0.2) (38); ii) there was evidence of a dose-response relation; iii) there was evidence that all plausible residual confounding could decrease the magnitude of the association (39). Severity factors were graded downward (very low) if there are risk of bias (36). Only severity factors with moderate or high-quality of evidence were included in the review.

## Results

### Studies selection

We identified a total of 9,983 titles through three database searches (PubMed = 6,910; Embase = 2,845; and Cochrane = 228). After title screening and abstract review 222 full text articles were assessed for eligibility, 105 were excluded, and 117 articles were finally included in this review (See PRISMA flow diagram in Figure 1).

**Figure 1.**
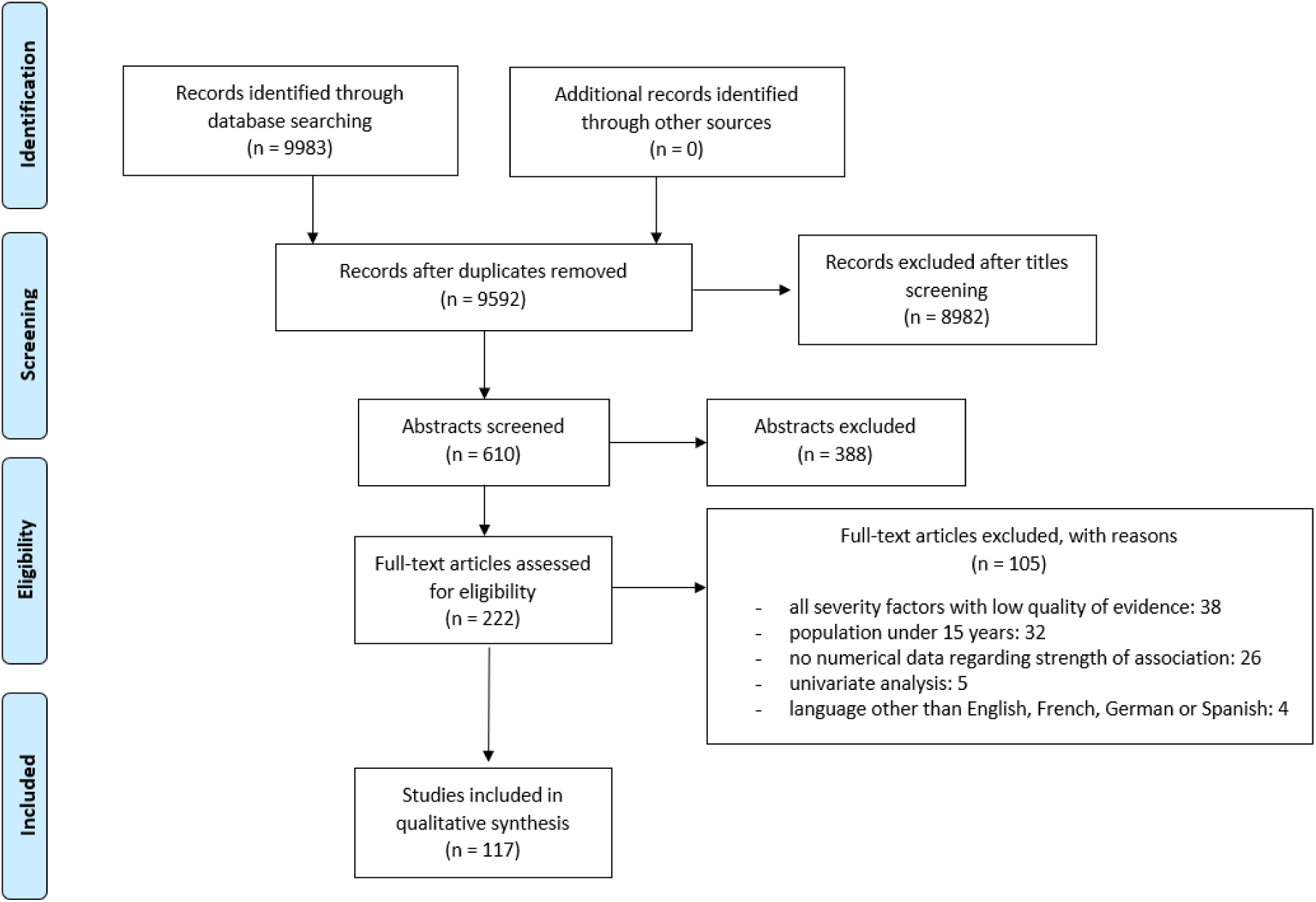
Selection of studies using the Preferred reporting item for systematic reviews and meta-analyses (PRISMA) flow diagram.

### Studies characteristics

Of the 117 studies included, 114 studies were conducted in 41 individual countries and 3 in multiple countries. The countries accounting for the highest number of studies were Ethiopia with 11 (9.4%) publications, and Brazil, China, and the United States with 9 (7.7%) publications each. Seventy-six (64.9%) studies were retrospective cohorts, 27 (23.1%) were prospective cohorts, 7 were case-controls studies, 6 were cross-sectionals, and 1 both retrospective and prospective. Participant follow-up duration in the studies ranged from 14 days to 3650 days with a median of 255.0 days (IQR, 180-360).

Mortality was reported in 98 (83.8%) studies, severe morbidity in 8 (6.8%), unsuccessful treatment (default, death or treatment failure) in 7 (6.0%), and sequelae (lung impairment) in 4 (3.4%).

### Studies population characteristics

The size of the study populations ranged from 44 to 2,551,058 with a median of 769 (IQR, 299- 3245) individuals; participants’ median age ranged from 28 to 80 years, and the proportion of male participants ranged from 10.4% to 100% with a median of 62.9% (IQR, 54.0%- 69.2%). Forty-three (36.8%) studies were conducted among inpatients, 10 (8.5%) among outpatients and 64 (54.7%) among both inpatients and outpatients. Ten (8.5%) studies did not specify HIV status. Thirty-nine (33.3%) studies were conducted among PLWH only, 4 (3.4%) among people without HIV only and 64 (54.70%) among both people with and without HIV. Among the latter, the proportion of PLWH ranged from 20.0% to 68.4% with a median of 44.2% (IQR, 20.0%- 68.4%). In studies including PLWH, median CD4 counts ranged from 25 to 290 cells/μL with a median of 96 cells/μL (IQR, 54-172). Proportion of patients with CD4 count <200 cells/μL ranged from 16.7% to 100% with a median of 50.1 % (IQR, 36.9 %- 60.6%).

Thirty-seven (31.6%) studies included only participants with pulmonary TB, 80 (68.4%) included participants with both pulmonary and extra pulmonary. Among these, the proportion of participants with pulmonary TB ranged from 15.2% to 95.5% with a median of 74.0% (IQR, 56.4%- 81.9%). The proportion of participants with confirmed TB ranged from 2.0% to 100% with a median of 64.0% (IQR, 41.3%-100%).

### Factors associated with TB severity

In studies that included only PLWH, 31 severity factors or groups of severity factors were reported as associated with mortality. Among them, six were also associated with severe morbidity and three with unsuccessful treatment. No factor associated with sequelae was reported (Table 1). Factors related to the host or mixed such as older age, anemia, immunosuppression, comorbidities, low BMI, fever, tachycardia, dyspnea, low blood pressure or social factors were the most frequently reported (72 times). Variables related to key disease characteristics such as bacillary load (smear positivity or high positivity level, and culture positivity) or disease dissemination (LAM positivity, extra pulmonary involvement, miliary or disseminated TB) were reported 14 times. No variable related to the extent of lung involvement on CXR was reported (Table 3). The strength of the association between severity factors and mortality is shown in figure 2 (40–77).

**Figure 2.**
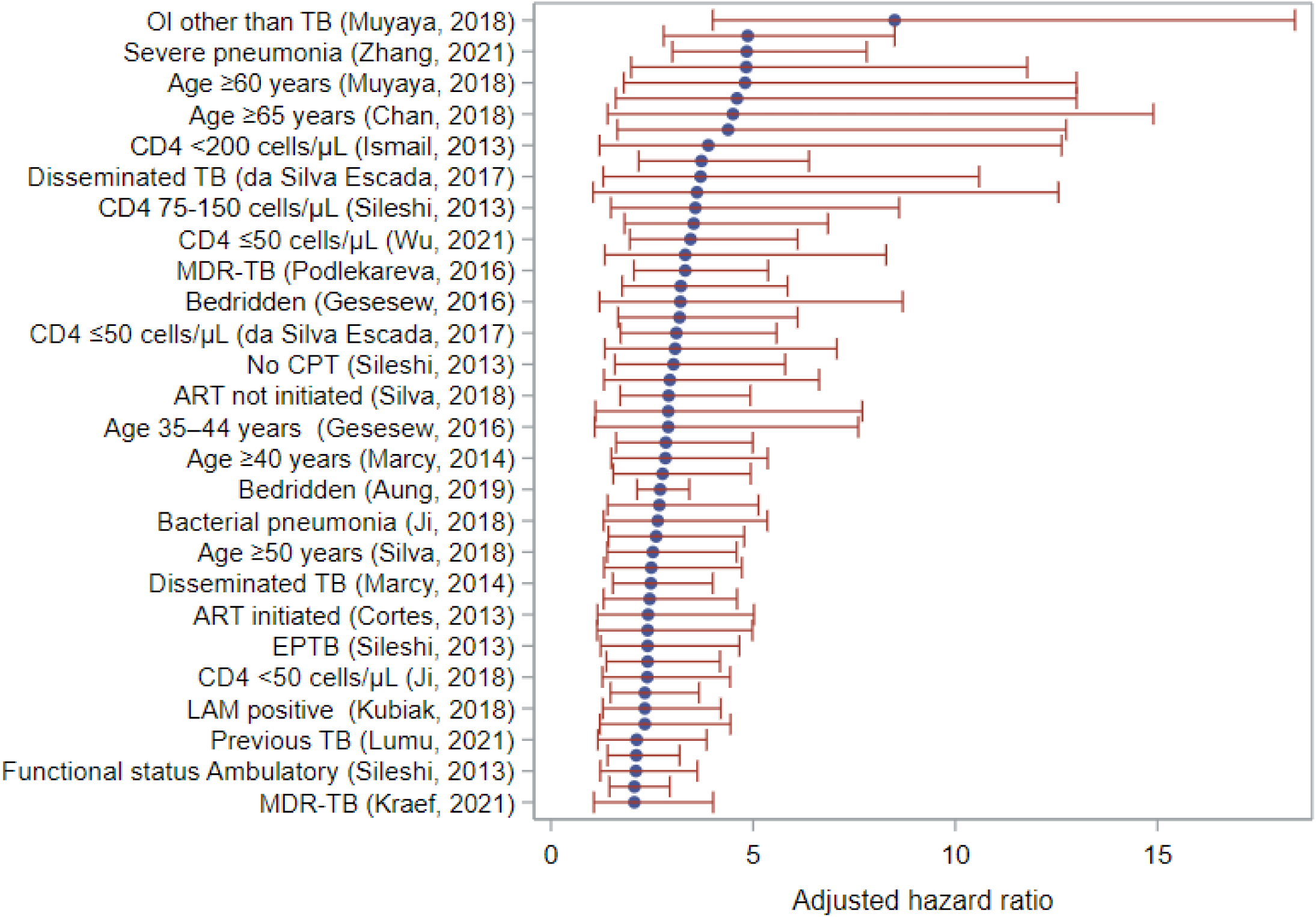
TB severity factors associated with mortality in PLWH: effect size was adjusted hazard ratio.

**Table 1.**
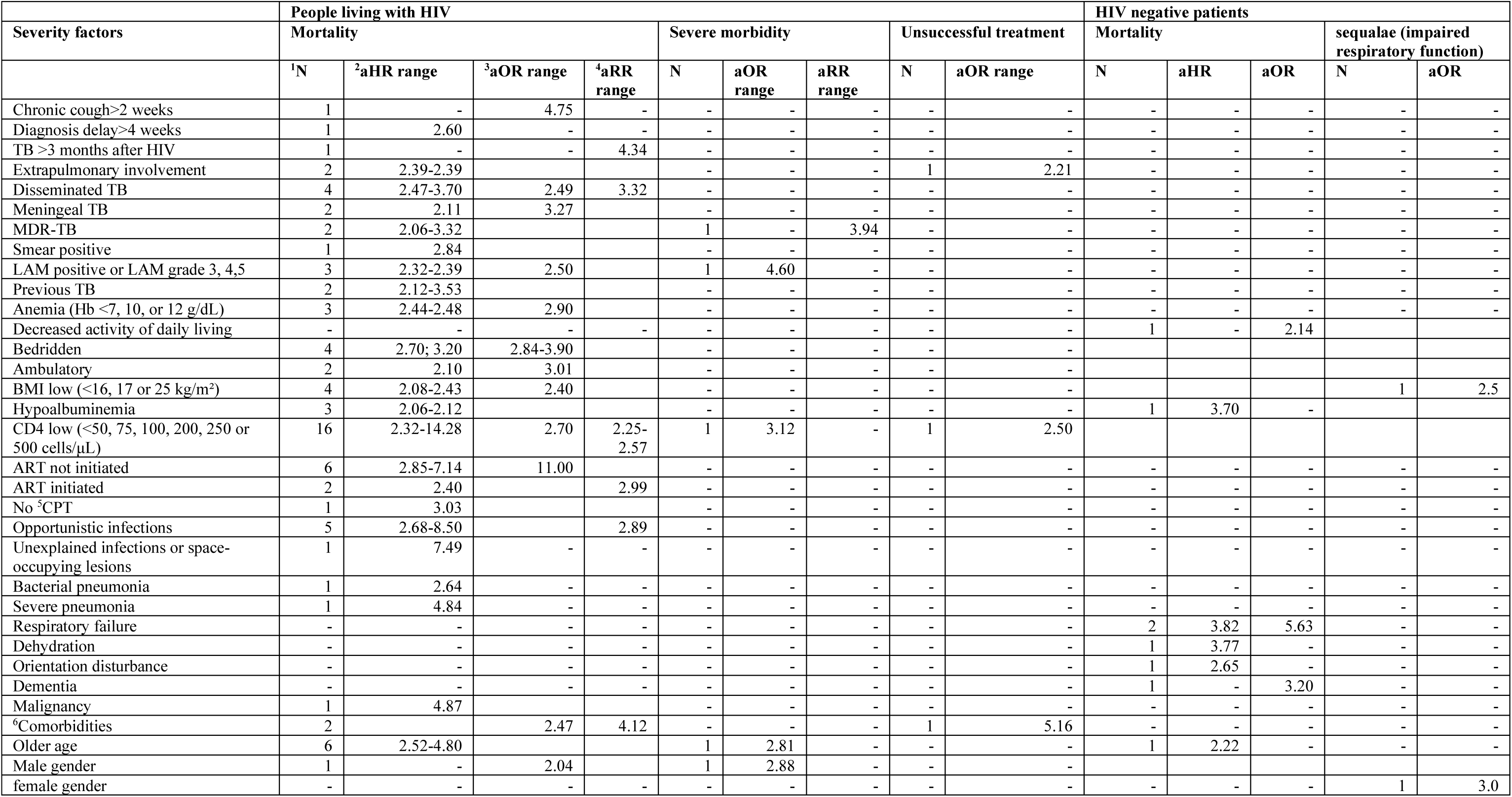
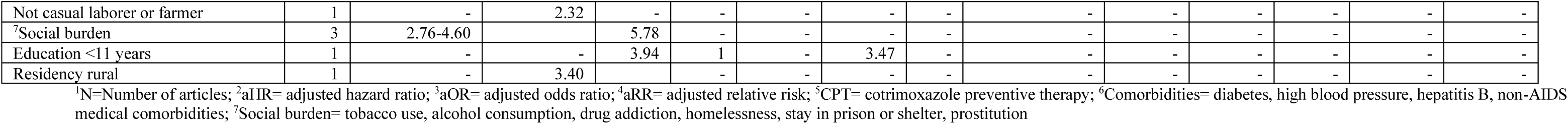
Severity factors in people living with HIV and in HIV negative patients.

| Severity factors | People living with HIV |  |  |  |  |  |  |  | HIV negative patients |  |  |  |  |  |
| --- | --- | --- | --- | --- | --- | --- | --- | --- | --- | --- | --- | --- | --- | --- |
|  | Mortality |  |  |  | Severe morbidity |  |  | Unsuccessful treatment |  | Mortality |  |  | sequalae (impaired respiratory function) |  |
|  | <sup>1</sup>N | <sup>2</sup>aHR range | <sup>3</sup>aOR range | <sup>4</sup>aRR range | N | aOR range | aRR range | N | aOR range | N | aHR | aOR | N | aOR |
| Chronic cough>2 weeks | 1 | - | 4.75 | - | - | - | - | - | - | - | - | - | - | - |
| Diagnosis delay>4 weeks | 1 | 2.60 | - | - | - | - | - | - | - | - | - | - | - | - |
| TB >3 months after HIV | 1 | - | - | 4.34 | - | - | - | - | - | - | - | - | - | - |
| Extrapulmonary involvement | 2 | 2.39-2.39 | - | - | - | - | - | 1 | 2.21 | - | - | - | - | - |
| Disseminated TB | 4 | 2.47-3.70 | 2.49 | 3.32 | - | - | - | - | - | - | - | - | - | - |
| Meningeal TB | 2 | 2.11 | 3.27 | - | - | - | - | - | - | - | - | - | - | - |
| MDR-TB | 2 | 2.06-3.32 | - | - | 1 | - | 3.94 | - | - | - | - | - | - | - |
| Smear positive | 1 | 2.84 | - | - | - | - | - | - | - | - | - | - | - | - |
| LAM positive or LAM grade 3, 4,5 | 3 | 2.32-2.39 | 2.50 | - | 1 | 4.60 | - | - | - | - | - | - | - | - |
| Previous TB | 2 | 2.12-3.53 | - | - | - | - | - | - | - | - | - | - | - | - |
| Anemia (Hb <7, 10, or 12 g/dL) | 3 | 2.44-2.48 | 2.90 | - | - | - | - | - | - | - | - | - | - | - |
| Decreased activity of daily living | - | - | - | - | - | - | - | - | - | 1 | - | 2.14 | - | - |
| Bedridden | 4 | 2.70; 3.20 | 2.84-3.90 | - | - | - | - | - | - | - | - | - | - | - |
| Ambulatory | 2 | 2.10 | 3.01 | - | - | - | - | - | - | - | - | - | - | - |
| BMI low (<16, 17 or 25 kg/m<sup>2</sup>) | 4 | 2.08-2.43 | 2.40 | - | - | - | - | - | - | - | - | - | 1 | 2.5 |
| Hypoalbuminemia | 3 | 2.06-2.12 | - | - | - | - | - | - | - | 1 | 3.70 | - | - | - |
| CD4 low (<50, 75, 100, 200, 250 or 500 cells/μL) | 16 | 2.32-14.28 | 2.70 | 2.25-2.57 | 1 | 3.12 | - | 1 | 2.50 | - | - | - | - | - |
| ART not initiated | 6 | 2.85-7.14 | 11.00 | - | - | - | - | - | - | - | - | - | - | - |
| ART initiated | 2 | 2.40 | - | 2.99 | - | - | - | - | - | - | - | - | - | - |
| No <sup>5</sup>CPT | 1 | 3.03 | - | - | - | - | - | - | - | - | - | - | - | - |
| Opportunistic infections | 5 | 2.68-8.50 | - | 2.89 | - | - | - | - | - | - | - | - | - | - |
| Unexplained infections or space-occupying lesions | 1 | 7.49 | - | - | - | - | - | - | - | - | - | - | - | - |
| Bacterial pneumonia | 1 | 2.64 | - | - | - | - | - | - | - | - | - | - | - | - |
| Severe pneumonia | 1 | 4.84 | - | - | - | - | - | - | - | - | - | - | - | - |
| Respiratory failure | - | - | - | - | - | - | - | - | - | 2 | 3.82 | 5.63 | - | - |
| Dehydration | - | - | - | - | - | - | - | - | - | 1 | 3.77 | - | - | - |
| Orientation disturbance | - | - | - | - | - | - | - | - | - | 1 | 2.65 | - | - | - |
| Dementia | - | - | - | - | - | - | - | - | - | 1 | - | 3.20 | - | - |
| Malignancy | 1 | 4.87 | - | - | - | - | - | - | - | - | - | - | - | - |
| <sup>6</sup>Comorbidities | 2 | - | 2.47 | 4.12 | - | - | - | 1 | 5.16 | - | - | - | - | - |
| Older age | 6 | 2.52-4.80 | - | - | 1 | 2.81 | - | - | - | 1 | 2.22 | - | - | - |
| Male gender | 1 | - | 2.04 | - | 1 | 2.88 | - | - | - | - | - | - | - | - |
| female gender | - | - | - | - | - | - | - | - | - | - | - | - | 1 | 3.0 |
| Not casual laborer or farmer | 1 | - | 2.32 | - | - | - | - | - | - | - | - | - | - | - |
| <sup>7</sup>Social burden | 3 | 2.76-4.60 |  | 5.78 | - | - | - | - | - | - | - | - | - | - |
| Education <11 years | 1 | - | - | 3.94 | 1 | - | 3.47 | - | - | - | - | - | - | - |
| Residency rural | 1 | - | 3.40 | - | - | - | - | - | - | - | - | - | - | - |

<sup>1</sup>N=Number of articles; <sup>2</sup>aHR= adjusted hazard ratio; <sup>3</sup>aOR= adjusted odds ratio; <sup>4</sup>aRR= adjusted relative risk; <sup>5</sup>CPT= cotrimoxazole preventive therapy; <sup>6</sup>Comorbidities= diabetes, high blood pressure, hepatitis B, non-AIDS medical comorbidities; <sup>7</sup>Social burden= tobacco use, alcohol consumption, drug addiction, homelessness, stay in prison or shelter, prostitution

**Table 2.** Severity factors in the general population.

| Severity factors | Mortality |  |  |  | Severe morbidity |  | Unsuccessful treatment |  |  |  | sequelae (impaired respiratory function) |  |  |
| --- | --- | --- | --- | --- | --- | --- | --- | --- | --- | --- | --- | --- | --- |
|  | <sup>1</sup>N | <sup>2</sup>aHR range | <sup>3</sup>aOR range | <sup>4</sup>aRR range | N | aOR range | N | aHR range | aOR range | aRR range | N | aOR range | aRR range |
| Possible TB | 1 | - | 6.28 | - | - | - | - | - | - | - | - | - | - |
| Probable TB | 1 | - | 3.38 | - | - | - | - | - | - | - | - | - | - |
| Bilateral lung involvement | 2 | - | 2.20-2.50 | - | - | - | - | - | - | - | - | - | - |
| Extrapulmonary involvement | 4 | 2.42 | 2.90-3.00 | - | - | - | - | - | - | - | - | - | - |
| <sup>5</sup>Severe extrapulmonary TB | 3 | - | 3.70-8.22 | - | - | - | - | - | - | - | - | - | - |
| Miliary TB | 2 | - | 2.38-3.79 | - | - | - | - | - | - | - | - | - | - |
| Clinical TB diagnosis | 1 | 4.96 | - | - | - | - | - | - | - | - | - | - | - |
| Previous TB | 3 | 2.90-3.48 | - | 2.20 | - | - | - | - | - | - | 1 | 4.3 | - |
| Smear or culture positive or bacillary load (1+ increase) | 6 | 2.23-3.07 | 2.49-2.50 | - | 1 | 3.76 | 1 | - | 3.37 | - | 1 | 3.5 | - |
| Smear negative | 2 | - | 2.40-2.70 | - | - | - | - | - | - | - | - | - | - |
| TB-specific abnormality on CXR | 1 | - | 2.84 | - | - | - | - | - | - | - | - | - | - |
| X-ray Unknown/not performed | 1 | - | 2.41 | - | - | - | - | - | - | - | - | - | - |
| Cavitation | 1 | - | 2.99 | - | - | - | 1 | - | 6.05 | - | - | - | - |
| No Cavitation | 1 | - | 5.26 | - | 1 | 3.22 | - | - | - | - | - | - | - |
| Infiltration > 2 lobes | - | - | - | - | 1 | 3.07 | - | - | - | - | - | - | - |
| No lobar consolidation | 1 | - | 20.00 | - | - | - | - | - | - | - | - | - | - |
| Fibrotic pattern | - | - | - | - | - | - | - | - | - | - | 1 | 3.5 | - |
| No productive cough | - | - | - | - | 1 | 2.77 | - | - | - | - | - | - | - |
| BMI low (<16 or 18.5 kg/m<sup>2</sup>) or weight loss | 6 | 2.53-4.54 | 2.19-3.90 | - | - | - | - | - | - | - | - | - | - |
| HIV positive | 24 | 2.51-7.70 | 2.29-8.08 | 2.32-4.48 | 1 | 5.74 | 1 | 2.85 | - | - | - | - | - |
| HIV status unknown | 3 | 2.17-2.80 | 3.51 | - | - | - | - | - | - | - | - | - | - |
| CD4 low (<50, 100, 200 or 350 cells/μL) | 3 | 6.31 | 4.27 | 2.18-10.29 | - | - | - | - | - | - | - | - | - |
| ART not initiated | 1 | - | 3.59 | - | - | - | - | - | - | - | - | - | - |
| <sup>6</sup>Other immunosuppression or Comorbidities | 15 | 2.12-4.10 | 2.13-7.00 | 4.90 | 1 | 5.74 | - | - | - | - | - | - | - |
| High blood urine nitrogen | 1 | - | 3.23 | - | - | - | - | - | - | - | - | - | - |
| Systolic blood pressure < 90 mmHg | 1 | - | 3.16 | - | - | - | - | - | - | - | - | - | - |
| Severe hypoxemia (P/F ratio < 100) | 2 | - | 4.30-4.70 | - | - | - | - | - | - | - | - | - | - |
| PaO<sub>2</sub>< 70 mmHg | 1 | - | 4.58 | - | - | - | - | - | - | - | - | - | - |
| SpO<sub>2</sub>< 90% | 1 | - | 4.25 | - | - | - | - | - | - | - | - | - | - |
| Respiratory Rate > 20 per minute | 1 | - | 3.29 | - | - | - | - | - | - | - | - | - | - |
| Dyspnea |  | - | - | - | 1 | 3.51 | - | - | - | - | - | - | - |
| Fever |  | - | - | - | 1 | 2.11 | - | - | - | - | - | - | - |
| Heart rate > 100 per minute | 1 | - | 2.27 | - | - | - | - | - | - | - | - | - | - |
| Septic shock | 1 | - | 5.21 | - | - | - | - | - | - | - | - | - | - |
| Anemia (Hb < 12 or 13 g/dL) |  | - | - | - | 1 | 2.91 | - | - | - | - | - | - | - |
| Hypoalbuminemia |  | - | - | - | 1 | 2.56 | - | - | - | - | - | - | - |
| Serum sodium, <135 mEq/L | 1 | - | 2.60 | - | - | - | - | - | - | - | - | - | - |
| Older age | 31 | 2.11-11.93 | 2.01-11.93 | 2.93-3.52 | - | - | - | - | - | - | - | - | - |
| Age < 50 years |  |  |  |  |  |  |  |  |  |  | 1 | 3.7 | - |
| Male gender | 1 | - | - | 5.50 | - | - | - | - | - | - | 1 |  | 5.1 |
| <sup>7</sup>Social burden | 2 |  | 2.80-2.88 | 3.30 |  |  | 1 |  |  | 3.58-4.67 | 1 |  | 6.7 |
| Education <11 years |  |  |  |  |  |  |  |  |  |  | 1 | 2.1 | - |
| Retirement | 1 | 2.40 | - | - | - | - | - | - | - | - |  |  | - |
| Long-term care facility | 2 | - | 2.10-2.36 | - | - | - | - | - | - | - |  |  | - |
| Married | 1 | 2.28 | - | - | - | - | - | - | - | - |  |  | - |
| Use of traditional medicine/herbs |  | - | - | - | 1 | 2.97 | - | - | - | - | - | - | - |
| No Physical activity, exercise |  | - | - | - | - | - | 1 | - | 5.96 | - | - | - | - |

<sup>1</sup>N=Number of articles; <sup>2</sup>aHR= adjusted hazard ratio; <sup>3</sup>aOR= adjusted odds ratio; <sup>4</sup>aRR= adjusted relative risk; <sup>5</sup>Severe extrapulmonary TB= meningitis, miliary, pericarditis, peritonitis, bilateral or extensive pleural effusion, spinal, genitourinary, and intestinal types; <sup>6</sup>Other immunosuppression or comorbidities= diabetes mellitus, malignancy, chronic kidney disease, chronic respiratory disease, hypertension, Ischemic heart disease, congestive heart failure, immunosuppressant use, liver failure or cirrhosis, cerebral vascular disease, cognitive impairment, pregnancy. <sup>7</sup>Social burden= tobacco use, alcohol consumption, drug addiction, homelessness, stay in prison or shelter, prostitution

**Table 3.** Summary of key findings.

| Severity factors | PLWH<br>n (n *HQE) | HIV-<br>n (n HQE) | Both<br>n (n HQE) |
| --- | --- | --- | --- |
| **Related to host or mixed**<br>(older age, anemia, immunosuppression, comorbidities, low BMI, fever, tachycardia, dyspnea, low blood pressure, social factors) | 72 (8) | 8 (2) | 92 (26) |
| **Bacillary load** (smear+, high positivity level, culture +) | 1 (0) | 0 | 9 (0) |
| **Extent of lung involvement** (cavitation, bilateral location, Infiltration > 2 lobes) | 0 | 0 | 8 (2) |
| **Disease dissemination** (LAM+, extra pulmonary involvement, miliary, disseminated TB) | 13 (0) | 0 | 9 (1) |

\*HQE= high quality of evidence

In studies enrolling only people without HIV, 9 severity factors were reported (Table 1). Among them, seven were associated with mortality and two with sequelae (impaired respiratory function). No factor was reported as associated with severe morbidity or unsuccessful treatment (Table 1). All factors were related to the host or mixed, and none was related to key disease characteristics (Table 3) (16,78–80).

In studies that included both PLWH and people without HIV, 47 severity factors or groups of severity factors were reported. Among them, 28 were associated only with mortality, seven only with severe morbidity, one only with unsuccessful treatment, three only with sequelae and eight with both mortality and at least one outcome among severe morbidity, unsuccessful treatment or sequalae (Table 2). Factors related to the host or mixed were the most frequently reported (92 times). Variables related to bacillary load, extent of lung involvement (cavitation, bilateral location, infiltration to more than 2 lobes) or disease dissemination were reported 26 times, 3 times with high quality of evidence (Table 3). The strength of the association between severity factors and mortality is shown in figure 3. (3,6,14,15,29,81–128).

**Figure 3.**
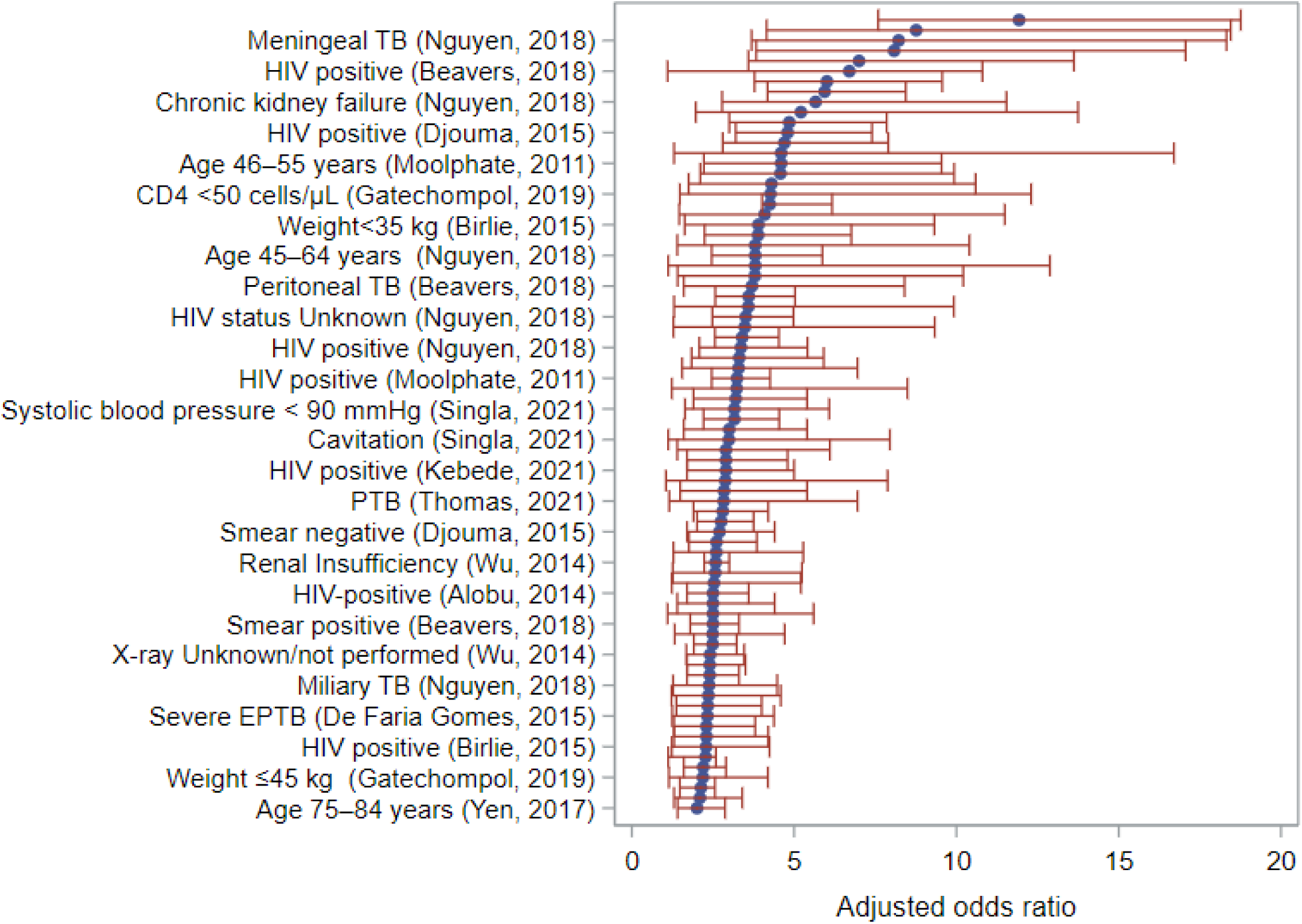
TB severity factors associated with mortality in the general population: effect size was adjusted odds ratio.

## Discussion

Our review enabled identifying multiple factors associated with TB severity. The most frequently reported severity factors were related to the host or mixed. However, there were also factors related to key TB disease characteristics such as bacillary load, extent of lung involvement and disease dissemination. These factors were reported 14 times in people with HIV and 26 times in studies that included both people with and without HIV (Table 3.).

The dominance of host-related and mixed factors as predictors of TB severity aligns with findings from prior systematic reviews. A review of mortality risk factors among TB patients in South Africa identified HIV infection and lower body weight as key contributors to TB mortality (129). Similarly, a meta-analysis of in-hospital mortality predictors found comorbid malignancy to be strongly associated with increased mortality risk (130). In critically ill TB patients, older age was identified as the primary factor influencing mortality (131). The relationship between older age, one of the most frequently reported factors, and TB severity is likely multifactorial, with older patients often presenting with weaker immune responses, delayed diagnosis, and many comorbidities.

Despite not being the most frequently reported factors, variables reflecting key disease characteristics bacillary load, extent of lung involvement and disease dissemination were nevertheless reported 40 times. Their frequency would likely have been much higher if their relationship with TB severity was systematically assessed in all studies, which was not the case as most studies only focused of host-related and mixed factors. This could explain why there is very little mention of these factors in previous reviews (129,131,132). Nonetheless, these factors highlight the intrinsic role of TB itself in determining disease severity, since they directly reflect the burden and the inherent aggressiveness and progression of *Mycobacterium tuberculosis* disease. Bacillary load, as measured by molecular assays, is not only a marker of disease severity but also an early predictor of treatment response, underscoring its importance in clinical management (30). Cavitations and extensive lung involvement are strong predictors of poor outcomes, directly linking the extent of bacterial activity and tissue damage to mortality and treatment failure (26). LAM positivity strongly correlates with early mortality in hospitalized patients, reflecting a systemic spread of TB in advanced disease stages (28). This is further supported by studies which found that miliary TB and extrapulmonary involvement significantly increased the risk of death (92,133). Disease-specific characteristics therefore not only reinforce the pathogenic role of TB but also provide actionable insights for clinical practice.

One limitation of this study is that, as a systematic review, it relies entirely on data from primary research, which may vary in quality and methodology. While the GRADE framework was used to assess quality of evidence, residual biases or errors in the original studies could influence the findings. Furthermore, there were no studies allowing to assess differences between PLWH and HIV negative patients. Besides, by restricting the review to studies published in English, French, German, and Spanish, important research published in other languages, particularly in high-burden countries like China, may have been excluded. This could reduce the comprehensiveness of the review.

Despite these limitations, a major strength of this review is its broad inclusion of studies from all designs, settings and regions, allowing for the exhaustive identification of severity factors reported by 117 studies, a number far higher than in most systematic reviews. The inclusion of studies from 41 countries and three international settings ensures a wide geographical representation; this diversity supports the applicability of the findings to both high-burden and low-burden countries. Besides, the use of the GRADE system classification ensures that the evidence is critically appraised for quality, enhancing the reliability of the findings.

## Conclusion

This review enabled identifying many factors associated with TB severity. Most factors were typically related to the host or both to the host and TB. However, the review also brought evidence that TB severity is related as well to bacillary load, extent of lung involvement and disease dissemination. It could be important to consider these factors in TB severity assessment and to assess them systematically in prospective studies. In addition, it may be useful to develop a specific TB severity score for people with HIV who have different and severe forms of TB, especially those who are severely immunosuppressed.

## List of abbreviations

TB: Tuberculosis
HIV: Human Immunodeficiency Virus
AIDS: Acquired Immunodeficiency Syndrome
PLWH: People Living With HIV
HQE: High Quality of Evidence
IRIS: Immune Reconstitution Inflammatory Syndrome
BMI: Body Mass Index
MUAC: Mid-Upper Arm Circumference
CXR: Chest X-ray
LAM: Lipoarabinomannan
ART: Antiretroviral Therapy
CPT: Cotrimoxazole Preventive Therapy
aHR: Adjusted Hazard Ratio
aOR: Adjusted Odds Ratio
aRR: Adjusted Relative Risk
PRISMA: Preferred Reporting Items for Systematic Reviews and Meta-Analyses
PROSPERO: Prospective Register of Systematic Reviews
RAND: Research and Development (used in RAND Appropriateness Method)
MeSH: Medical Subject Headings
MDR-TB: Multidrug-Resistant Tuberculosis
RR: Relative Risk
PaO2: Partial Pressure of Oxygen
SpO2: Peripheral Oxygen Saturation

## Declarations

Ethics approval and consent to participate.

Not applicable.

## Consent for publication

Not applicable.

## Availability of data and materials

Data and materials are available from the corresponding author on reasonable request.

## Authors’ contributions

**Robert Akpata**: developed search equations in the different databases, completed the screening of articles by titles, independently reviewed abstracts for eligibility criteria, extracted data from the selected publications, and wrote the first draft.

**Anani Badjé**: independently reviewed abstracts for eligibility criteria, reviewed and validated data extracted from the selected publications, and reviewed the first draft.

**Jean-Baptiste Ntakpé**: independently reviewed abstracts for eligibility criteria, reviewed and validated data extracted from the selected publications.

**Angelo Attinsounon**: independently reviewed abstracts for eligibility criteria.

**Nathalie de Castro**: reviewed the first draft.

**Didier Laureillard**: reviewed the first draft.

**Marcel Zannou**: reviewed the first draft.

**Olivier Marcy**: independently reviewed abstracts for eligibility criteria, reviewed and validated data extracted from the selected publications, and reviewed the first draft.

## Data Availability

All data produced in the present work are contained in the manuscript

## Acknowledgements

We express our profound gratitude to Coralie Thore, the librarian of the University of Bordeaux who validated the search equations; the panel of TB experts who helped categorize the severity factors and to Mélanie Plazy for her very helpful advice.

