## Supplementary appendix for "Factors associated with severity of pulmonary tuberculosis in people living with and without HIV: a systematic review"

#### Appendix 1. Keywords and MeSH terms for search equations

| Concept | Keywords | MeSH Terms |
|---|---|---|
| Severity | Seriousness OR serious OR severity OR severe OR risk OR life-threatening | Risk OR Risk Factors OR Catastrophic Illness |
| Morbidity | Morbidity OR complications OR hospitalization OR hospitalized OR inpatient OR critical OR paradoxical reaction OR immune reconstitution OR IRIS OR failure OR failed OR worsen | Morbidity OR hospitalization OR inpatients OR critical care OR Treatment Failure |
| Sequelae | Sequelae OR sequels OR impairment OR disorder OR deficit OR disability OR dysfunction OR disturbance | Sequelae OR impairment OR disorder OR deficit OR disability OR dysfunction OR disturbance |
| Deaths | Death OR dead OR mortality OR deceased | Death |
| Tuberculosis | Tuberculosis OR TB OR Mycobacterium tuberculosis OR Koch | Tuberculosis |
| HIV | HIV OR AIDS OR human immunodeficiency virus OR acquired immunodeficiency syndrome | HIV infections |

| | | |
|---|---|---|
| Grading systems | Score OR scale OR index OR assessment OR grade OR predict OR predictive OR prognosis OR measure OR factor OR risk | Severity of Illness Index OR Early Warning Score OR Patient Outcome Assessment OR prognosis |

### Appendix 2. Search equations

The search equations in PubMed were:

i) factors associated with TB severity: (Tubercul*[Title] OR TB[Title] OR koch*[Title] OR tuberculosis[MeSH Major Topic]) AND (Seriou*[Title] OR sever*[Title] OR risk*[Title] OR life-threatening[Title] OR complicat*[Title] OR morbid*[Title] OR hospital*[Title] OR inpatient*[Title] OR intensive [Title] OR critical*[Title] OR paradoxical reaction[Title] OR IRIS [Title] OR immune reconstitution [Title] OR fail*[Title] OR worse* [Title] OR sequel*[Title] OR impair*[Title] OR disorders[Title] OR deficit[Title] OR Disability[Title] OR dysfunction[Title] OR disturbance [Title] OR Death[Title] OR death [Title] OR mortality [Title] OR deceased [Title] OR risk [MeSH Major Topic] OR Risk Factors [MeSH Major Topic] OR Catastrophic Illness [MeSH Major Topic] OR Morbidity [MeSH Major Topic] OR hospitalization [MeSH Major Topic] OR inpatients [MeSH Major Topic] OR critical care [MeSH Major Topic] OR Treatment Failure [MeSH Major Topic] OR Neurologic Manifestations [MeSH Major Topic] OR Death [MeSH Major Topic]**)** ;

ii) previously developed TB severity grading systems: (Tuberculosis[Title] OR TB[Title] OR koch*[Title] OR tuberculosis[MeSH Major Topic]) AND (Score[Title] OR scale[Title] OR index[Title] OR assess*[Title] OR grade[Title] OR predict* [Title] OR prognos*[Title] OR measure* [Title] OR Severity of Illness Index [MeSH Major Topic] OR Early Warning Score[MeSH Major Topic] OR Patient Outcome Assessment[MeSH Major Topic] OR prognosis[MeSH Major Topic]) AND (Seriou*[Title] OR sever*[Title] OR risk*[Title] OR life-threatening[Title] OR complicat*[Title] OR morbid*[Title] OR hospital*[Title] OR inpatient*[Title] OR intensive [Title] OR critical*[Title] OR paradoxical reaction[Title] OR IRIS [Title] OR immune reconstitution [Title] OR fail*[Title] OR worse* [Title] OR sequel*[Title] OR impair*[Title] OR disorders[Title] OR deficit[Title] OR Disability[Title] OR dysfunction[Title] OR disturbance [Title]

OR Death[Title] OR death [Title] OR mortality [Title] OR deceased [Title] OR risk [MeSH Major Topic] OR Risk Factors [MeSH Major Topic] OR Catastrophic Illness [MeSH Major Topic] OR Morbidity [MeSH Major Topic] OR hospitalization [MeSH Major Topic] OR inpatients [MeSH Major Topic] OR critical care [MeSH Major Topic] OR Treatment Failure [MeSH Major Topic] OR Neurologic Manifestations [MeSH Major Topic] OR Death [MeSH Major Topic])

### Appendix 3. List of studies and severity factors included

| Author | Year | Country | Size | HIV_status | Outcome | Severty_factor | Effect_measure | Effect_size | Lower_CI | Upper_CI | Reference_category | Quality_of_Evidence |
|---|---|---|---|---|---|---|---|---|---|---|---|---|
| Adamu | 2017 | Nigeria | 1424 | Mixed | Mortality | Clinical TB diagnosis | aHR | 4.96 | 2.69 | 9.17 | Bacteriological TB diagnosis | moderate (3+) |
| Adamu | 2017 | Nigeria | 1424 | Mixed | Mortality | Previous TB | aHR | 3.48 | 2.54 | 4.77 | No Previous TB treatment | moderate (3+) |
| Adamu | 2017 | Nigeria | 1424 | Mixed | Mortality | Age 45–54 years | aHR | 2.37 | 1.44 | 3.92 | Age group 15-24 years | moderate (3+) |
| Alobu | 2014 | Nigeria | 1668 | Mixed | Mortality | EPTB | aOR | 2.90 | 1.40 | 6.10 | Pulmonary TB | moderate (3+) |
| Alobu | 2014 | Nigeria | 1668 | Mixed | Mortality | HIV-positive | aOR | 2.50 | 1.70 | 3.60 | HIV-negative | moderate (3+) |
| Alobu | 2014 | Nigeria | 1668 | Mixed | Mortality | Smear negative | aOR | 2.40 | 1.70 | 3.50 | Smear-positive PTB | moderate (3+) |
| Alvarez-Uria | 2016 | India | 1615 | Positive | Mortality | Albumin (1 g/dL increase) | aHR | 0.47 | 0.41 | 0.54 | NA | high (4+) |
| Aung | 2019 | Myanmar | 3598 | Positive | Mortality | Bedridden | aHR | 2.70 | 2.13 | 3.42 | Not bedridden | moderate (3+) |
| Ballayira | 2021 | Mali | 1362 | Mixed | Mortality | HIV positive | aHR | 7.10 | 3.53 | 14.26 | HIV negative status | high (4+) |
| Ballayira | 2021 | Mali | 1362 | Mixed | Mortality | Age ≥45 years | aHR | 2.16 | 1.40 | 3.33 | Age<45 years old | moderate (3+) |

| | | | | | | | | | | | | |
|---|---|---|---|---|---|---|---|---|---|---|---|---|
| Ballayira | 2021 | Mali | 1362 | Mixed | Mortality | Initial weight<40kg | aHR | 4.06 | 2.42 | 6.81 | Initial weight≥40kg | moderate (3+) |
| Bastos | 2016 | Portugal | 681 | Mixed | Mortality | Age ≥50 years | aOR | 2.90 | 1.70 | 4.80 | Age<50 years old | moderate (3+) |
| Bastos | 2016 | Portugal | 681 | Mixed | Mortality | Hypoxemic respiratory failure | aOR | 4.70 | 2.80 | 7.90 | No hypoxemic respiratory failure | moderate (3+) |
| Bastos | 2016 | Portugal | 681 | Mixed | Mortality | Bilateral lung involvement | aOR | 2.50 | 1.40 | 4.40 | Unilateral lung involvement | moderate (3+) |
| Bastos | 2016 | Portugal | 681 | Mixed | Mortality | ≥ 1 significant comorbidity | aOR | 2.30 | 1.30 | 3.80 | No significant comorbidity | moderate (3+) |
| Beavers | 2018 | United States | 1304 cases and 1,039 controls | Mixed | Mortality | HIV positive | aOR | 6.70 | 1.10 | 10.80 | HIV negative status | high (4+) |
| Beavers | 2018 | United States | 1304 cases and 1,039 controls | Mixed | Mortality | Peritoneal TB | aOR | 3.70 | 1.60 | 8.40 | No peritoneal TB | moderate (3+) |
| Beavers | 2018 | United States | 1304 cases and 1,039 controls | Mixed | Mortality | Cancer | aOR | 3.20 | 1.90 | 5.40 | No cancer | moderate (3+) |
| Beavers | 2018 | United States | 1304 cases and 1,039 controls | Mixed | Mortality | Cognitive impairment | aOR | 2.80 | 1.90 | 4.20 | No cognitive impairment | moderate (3+) |
| Beavers | 2018 | United States | 1304 cases and 1,039 controls | Mixed | Mortality | Immunosuppressive medications | aOR | 2.50 | 1.10 | 5.60 | No immunosuppressive medications | moderate (3+) |

| | | | | | | | | | | | | |
|---|---|---|---|---|---|---|---|---|---|---|---|---|
| Beavers | 2018 | United States | 1304 cases and 1,039 controls | Mixed | Mortality | Smear positive | aOR | 2.50 | 1.80 | 3.30 | Smear negative | moderate (3+) |
| Beavers | 2018 | United States | 1304 cases and 1,039 controls | Mixed | Mortality | COPD | aOR | 2.40 | 1.70 | 3.30 | No chronic obstructive pulmonary disease | moderate (3+) |
| Beavers | 2018 | United States | 1304 cases and 1,039 controls | Mixed | Mortality | Bilateral lung involvement | aOR | 2.20 | 1.60 | 2.90 | Unilateral disease | moderate (3+) |
| Beavers | 2018 | United States | 1304 cases and 1,039 controls | Mixed | Mortality | Diagnosed in long-term care facility | aOR | 2.10 | 1.30 | 3.40 | Not diagnosed in long-term care facility | moderate (3+) |
| Bhurayanontachai | 2016 | Thailand | 268 | Mixed | Significant morbidity | Sputum smear (1+ increase) | aOR | 3.76 | 1.90 | 7.47 | NA | moderate (3+) |
| Bhurayanontachai | 2016 | Thailand | 268 | Mixed | Significant morbidity | Dyspnea | aOR | 3.51 | 2.02 | 6.11 | No dyspnea | moderate (3+) |
| Bhurayanontachai | 2016 | Thailand | 268 | Mixed | Significant morbidity | Infiltration > 2 lobes | aOR | 3.07 | 1.78 | 5.29 | Extension of infiltration less than 2 lobes on chest X-ray | moderate (3+) |
| Bhurayanontachai | 2016 | Thailand | 268 | Mixed | Significant morbidity | Fever | aOR | 2.11 | 1.20 | 3.71 | No fever | moderate (3+) |
| Bhurayanontachai | 2016 | Thailand | 268 | Mixed | Significant morbidity | Albumin (1 g/dL increase) | aOR | 0.39 | 0.26 | 0.59 | NA | moderate (3+) |
| Bhurayanontachai | 2016 | Thailand | 268 | Mixed | Significant morbidity | Productive cough | aOR | 0.36 | 0.20 | 0.64 | No productive cough | moderate (3+) |

| | | | | | | | | | | | | |
|---|---|---|---|---|---|---|---|---|---|---|---|---|
| Bhurayanontachai | 2016 | Thailand | 268 | Mixed | Significant morbidity | Cavitation | aOR | 0.31 | 0.15 | 0.66 | No cavity on chest X-ray | moderate (3+) |
| Bigna | 2015 | Cameroon | 99 | Positive | Mortality | non-AIDS comorbidity | aOR | 2.47 | 1.22 | 8.02 | Absence of non-AIDS comorbidity | moderate (3+) |
| Birlie | 2015 | Ethiopia | 810 | Mixed | Mortality | Weight<35 kg | aOR | 3.90 | 1.63 | 9.32 | Age≥35 | moderate (3+) |
| Birlie | 2015 | Ethiopia | 810 | Mixed | Mortality | HIV positive | aOR | 2.29 | 1.23 | 4.24 | HIV negative status | moderate (3+) |
| Bouton | 2019 | Ghana | 394 | Mixed | Mortality | HIV positive | aHR | 2.69 | 1.51 | 4.80 | HIV negative status | moderate (3+) |
| Bukundi | 2021 | Tanzania | 53753 | Mixed | Mortality | HIV positive | aHR | 2.51 | 2.26 | 2.79 | HIV negative status | moderate (3+) |
| Camara | 2017 | Guinea | 573 | Positive | Mortality | Comorbidities | RR | 4.12 | 2.10 | 8.10 | No comorbidities (diabetes, high blood pressure or hepatitis B) | moderate (3+) |
| Camara | 2017 | Guinea | 573 | Positive | Mortality | ≥2 opportunistic infections | RR | 2.89 | 1.39 | 6.02 | 1 opportunistic infection besides TB | moderate (3+) |
| Camara | 2017 | Guinea | 573 | Positive | Mortality | CD4 ≤200 cells/µL | RR | 2.25 | 1.16 | 4.37 | CD4>200 | moderate (3+) |
| Carroll | 2020 | Myanmar | 1344 | Mixed | Mortality | Age ≥55 years | RR | 3.52 | 2.11 | 5.85 | 16-34 years | moderate (3+) |
| Carroll | 2020 | Myanmar | 1344 | Mixed | Mortality | HIV Positive on ART | RR | 2.32 | 1.42 | 3.82 | HIV negative status | moderate (3+) |
| Chan | 2018 | Hong Kong SAR, China | 299 | Positive | Mortality | Drug addiction | aHR | 4.60 | 1.60 | 13.00 | No history of drug addiction | moderate (3+) |
| Chan | 2018 | Hong Kong SAR, China | 299 | Positive | Mortality | Age ≥65 years | aHR | 4.50 | 1.40 | 14.90 | Age < 65 years | moderate (3+) |

| | | | | | | | | | | | | |
|---|---|---|---|---|---|---|---|---|---|---|---|---|
| Chan | 2018 | Hong Kong SAR, China | 299 | Positive | Mortality | CD4 <50 cells/μL | aHR | 2.90 | 1.10 | 7.70 | CD4 cell count ≥50 | moderate (3+) |
| Charoensakulchai | 2021 | Thailand | 487 | Mixed | Unsuccessful treatment | Unemployment | aHR | 3.12 | 1.41 | 6.86 | Others occupation | moderate (3+) |
| Charoensakulchai | 2021 | Thailand | 487 | Mixed | Unsuccessful treatment | HIV positive | aHR | 2.85 | 1.25 | 6.46 | HIV negative status | moderate (3+) |
| Chushkin | 2017 | Russian Federation | 214 | Unknown | Sequelae | Age < 50 years | aOR | 3.77 | 1.95 | 7.25 | Age ≥ 50 years | moderate (3+) |
| Chushkin | 2017 | Russian Federation | 214 | Unknown | Sequelae | Previous TB | aOR | 4.33 | 1.86 | 10.11 | No recurrence of tuberculosis | moderate (3+) |
| Chushkin | 2017 | Russian Federation | 214 | Unknown | Sequelae | Positive culture in the past | aOR | 3.50 | 1.72 | 7.11 | No positive culture in the past | moderate (3+) |
| Chushkin | 2017 | Russian Federation | 214 | Unknown | Sequelae | High school education only | aOR | 2.11 | 1.03 | 4.33 | College education | moderate (3+) |
| Conesa-Botella | 2011 | Uganda | 90 | Positive | Significant morbidity | ART not initiated and LAM positive | aOR | 4.60 | 1.50 | 13.80 | Pre-HAART LAM negative | moderate (3+) |
| Cortes | 2013 | International | 253 | Positive | Mortality | ART initiated | aHR | 2.40 | 1.15 | 5.02 | ART after TB diagnosis | moderate (3+) |
| da Silva Escada | 2017 | Brazil | 310 | Positive | Mortality | Disseminated TB | aHR | 3.70 | 1.29 | 10.59 | Extrapulmonary, localized TB | moderate (3+) |
| da Silva Escada | 2017 | Brazil | 310 | Positive | Mortality | CD4 ≤50 cells/μL | aHR | 3.10 | 1.72 | 5.58 | CD4> 50 | moderate (3+) |
| De Faria Gomes | 2015 | Brazil | 1257 | Mixed | Mortality | Age ≥40 years | aOR | 2.61 | 1.76 | 3.85 | Age<40 years | moderate (3+) |
| De Faria Gomes | 2015 | Brazil | 1257 | Mixed | Mortality | HIV positive | aOR | 6.01 | 3.78 | 9.56 | HIV negative status | high (4+) |

| | | | | | | | | | | | | |
|---|---|---|---|---|---|---|---|---|---|---|---|---|
| De Faria Gomes | 2015 | Brazil | 1257 | Mixed | Mortality | Severe EPTB | aOR | 2.33 | 1.24 | 4.38 | No severe extrapulmonary TB | moderate (3+) |
| de Mendonça | 2021 | Brazil | 258 | Mixed | Significant morbidity | Anemia (Hb <13 g/dL or <12 g/dL) | aOR | 2.91 | 1.16 | 7.29 | No Anemia ( | moderate (3+) |
| Deribe | 2015 | Ethiopia | 276 | Positive | Mortality | Chronic cough>2 weeks | aOR | 4.75 | 2.14 | 10.56 | No chronic cough | moderate (3+) |
| Deribe | 2015 | Ethiopia | 276 (69 cases: TB-HIV-coinfected patients who died and 207 controls: TB-HIV-coinfected patients who were alive) | Positive | Mortality | Functional status Ambulatory | aOR | 3.01 | 1.30 | 6.98 | Functional status: working | moderate (3+) |
| Deribe | 2015 | Ethiopia | 276 (69 cases: TB-HIV-coinfected patients who died and 207 controls: TB-HIV-coinfected | Positive | Mortality | Bedridden | aOR | 2.84 | 1.17 | 6.89 | Functional status: working | moderate (3+) |

| | | | | | | | | | | | | |
|---|---|---|---|---|---|---|---|---|---|---|---|---|
| | | | patients who were alive) | | | | | | | | | |
| Deribe | 2015 | Ethiopia | 276 (69 cases: TB-HIV-coinfected patients who died and 207 controls: TB-HIV-coinfected patients who were alive) | Positive | Mortality | Male gender | aOR | 2.04 | 1.04 | 4.02 | Female gender | moderate (3+) |
| Djouma | 2015 | Cameroon | 3245 (362 cases of death and 2883 patients successfully treated) | Mixed | Mortality | HIV positive | aOR | 4.80 | 3.20 | 7.40 | HIV negative status | moderate (3+) |
| Djouma | 2015 | Cameroon | 3245 (362 cases of death and 2883 patients successfully treated) | Mixed | Mortality | EPTB | aOR | 3.00 | 1.60 | 5.40 | Sputum smear positive pulmonary TB | moderate (3+) |

| | | | | | | | | | | | | |
|---|---|---|---|---|---|---|---|---|---|---|---|---|
| Djouma | 2015 | Cameroon | 3245 (362 cases of death and 2883 patients successfully treated) | Mixed | Mortality | Smear negative | aOR | 2.70 | 1.70 | 4.40 | Sputum smear positive pulmonary TB | moderate (3+) |
| Elhidsi | 2021 | Indonesia | 111 | Mixed | Mortality | Severe hypoxemia (P/F ratio < 100) | aOR | 4.30 | 1.75 | 10.59 | Mild-moderate hypoxemia | moderate (3+) |
| Elhidsi | 2021 | Indonesia | 111 | Mixed | Mortality | Renal Insufficiency | aOR | 4.09 | 1.46 | 11.49 | No renal Insufficiency | moderate (3+) |
| Engelbrecht | 2017 | South Africa | 40813 | Positive | Unsuccessful treatment | CD4 ≥351 cells/μL | aOR | 0.40 | 0.36 | 0.44 | CD4 1-201 | moderate (3+) |
| Ferreira | 2018 | Brazil | 120 | Positive | Mortality | CD4 >200 cells/μL | aHR | 0.26 | 0.08 | 0.87 | CD4 count < 200 | moderate (3+) |
| Ferreira | 2018 | Brazil | 120 | Positive | Mortality | Normal albuminemia | aHR | 0.47 | 0.26 | 0.85 | Hypoalbuminemia | moderate (3+) |
| Field | 2014 | South Africa | 3170 | Mixed | Mortality | Possible TB | RR | 6.28 | 3.18 | 12.43 | Confirmed TB | high (4+) |
| Field | 2014 | South Africa | 3170 | Mixed | Mortality | Probable TB | RR | 3.38 | 1.65 | 6.93 | Confirmed TB | moderate (3+) |
| Field | 2014 | South Africa | 3170 | Mixed | Mortality | HIV Positive not on ART | RR | 3.59 | 1.94 | 6.66 | Negative/unknown HIV/ART status | moderate (3+) |
| Field | 2014 | South Africa | 3170 | Mixed | Mortality | HIV Positive on ART | RR | 3.17 | 1.56 | 6.44 | Negative/unknown HIV/ART status | moderate (3+) |
| Field | 2014 | South Africa | 3170 | Mixed | Mortality | > = 1 previous TB episode | RR | 2.20 | 1.49 | 3.26 | First TB episode | moderate (3+) |

| | | | | | | | | | | | | |
|---|---|---|---|---|---|---|---|---|---|---|---|---|
| Fiogbe | 2019 | Benin | 189 | Negative | Sequelae | Female gender | aOR | 2.98 | 1.40 | 6.34 | Male gender | moderate (3+) |
| Fiogbe | 2019 | Benin | 189 | Negative | Sequelae | BMI >25 kg/m² | aOR | 0.40 | 0.26 | 0.52 | Normal weight (18 ,BMI ,25 kg/m2) | moderate (3+) |
| Gatechompol | 2019 | Thailand | 996 | Mixed | Mortality | Age ≥50 years | aOR | 3.89 | 2.24 | 6.76 | Age ≤50 years | moderate (3+) |
| Gatechompol | 2019 | Thailand | 996 | Mixed | Mortality | Weight ≤45 kg | aOR | 2.19 | 1.14 | 4.19 | Weight >45 kg | moderate (3+) |
| Gatechompol | 2019 | Thailand | 996 | Mixed | Mortality | ART not initiated | aOR | 3.59 | 1.30 | 9.92 | ART | moderate (3+) |
| Gatechompol | 2019 | Thailand | 996 | Mixed | Mortality | HIV positive | aOR | 3.31 | 1.84 | 5.91 | HIV negative status | moderate (3+) |
| Gatechompol | 2019 | Thailand | 996 | Mixed | Mortality | CD4 <50 cells/µL | aOR | 4.27 | 1.48 | 12.30 | CD4 cell count ≥50 | moderate (3+) |
| Gesesew | 2016 | Ethiopia | 272 | Positive | Mortality | Bedridden | aOR | 3.90 | 1.50 | 10.30 | Functional status Work | moderate (3+) |
| Gesesew | 2016 | Ethiopia | 272 | Positive | Mortality | Residency rural | aOR | 3.40 | 1.40 | 8.40 | Residence urban | moderate (3+) |
| Gesesew | 2016 | Ethiopia | 272 | Positive | Mortality | Bedridden | aHR | 3.20 | 1.20 | 8.70 | Functional status Work | moderate (3+) |
| Gesesew | 2016 | Ethiopia | 272 | Positive | Mortality | Age 35–44 years | aHR | 2.90 | 1.08 | 7.60 | 15–24 years | moderate (3+) |
| Gesesew | 2016 | Ethiopia | 272 | Positive | Mortality | WHO stage 2 | aHR | 0.20 | 0.06 | 0.50 | WHO stage 1 | moderate (3+) |
| Goto | 2018 | Japan | 367 | Negative | Mortality | Albumin (1 g/dL increase) | aHR | 0.27 | 0.16 | 0.46 | NA | high (4+) |
| Henegar | 2012 | Congo, Dem. Rep. | 5685 | Mixed | Mortality | EPTB | aHR | 2.42 | 1.52 | 3.85 | HIV negative Pulmonary tuberculosis smear-positive during intensive treatment phase | moderate (3+) |

| | | | | | | | | | | | | |
|---|---|---|---|---|---|---|---|---|---|---|---|---|
| Huerga | 2021 | Malawi | 387 | Positive | Mortality | Anemia (Hb <12 g/dL) | aOR | 2.90 | 1.40 | 5.70 | Hemoglobin ≥12 gr/dL | moderate (3+) |
| Huerga | 2021 | Malawi | 387 | Positive | Mortality | LAM positive | aOR | 2.50 | 1.10 | 5.80 | LAM negative | moderate (3+) |
| Huerga | 2021 | Malawi | 387 | Positive | Mortality | CD4 <200 cells/µL | aOR | 2.70 | 1.50 | 4.80 | CD4 ≥200 cells/µL | moderate (3+) |
| Huerga | 2021 | Malawi | 387 | Positive | Mortality | BMI <17 kg/m² | aOR | 2.40 | 1.30 | 4.30 | Body mass index ≥17 kg/m2 | moderate (3+) |
| Ismail | 2013 | Malaysia | 227 | Positive | Mortality | ≥3 Opportunistic infections | aHR | 3.61 | 1.04 | 12.55 | No Opportunistic infections | moderate (3+) |
| Ismail | 2013 | Malaysia | 227 | Positive | Mortality | 2 Opportunistic infections | aHR | 3.32 | 1.33 | 8.29 | No Opportunistic infections | moderate (3+) |
| Ismail | 2013 | Malaysia | 227 | Positive | Mortality | No Anti-retroviral therapy | aHR | 3.21 | 1.76 | 5.85 | Anti-retroviral therapy | moderate (3+) |
| Ismail | 2013 | Malaysia | 227 | Positive | Mortality | CD4 <200 cells/µL | aHR | 3.89 | 1.20 | 12.63 | ≥ 200 cells/µl | moderate (3+) |
| Ismail | 2013 | Malaysia | 227 | Positive | Mortality | 1 Opportunistic infection | aHR | 2.68 | 1.40 | 5.13 | No Opportunistic infections | moderate (3+) |
| Izudi | 2020 | Uganda | 1123 | Mixed | Mortality | Age ≥50 years | RR | 2.93 | 1.74 | 4.92 | Age 15–34 years | moderate (3+) |
| Izudi | 2020 | Uganda | 1123 | Mixed | Mortality | HIV positive | RR | 4.48 | 2.95 | 6.79 | HIV negative | moderate (3+) |
| Jacob | 2013 | Uganda | 368 | Positive | Significant morbidity | CD4 (100 cells/µL increase) | aOR | 0.32 | 0.17 | 0.60 | NA | moderate (3+) |
| Jacob | 2013 | Uganda | 368 | Positive | Significant morbidity | Male gender | aOR | 2.88 | 1.40 | 5.91 | Female | moderate (3+) |
| Ji | 2018 | China | 359 | Positive | Mortality | Age ≥60 years | aHR | 3.18 | 1.66 | 6.10 | Age ≤60 | moderate (3+) |

| | | | | | | | | | | | | |
|---|---|---|---|---|---|---|---|---|---|---|---|---|
| Ji | 2018 | China | 359 | Positive | Mortality | Bacterial pneumonia | aHR | 2.64 | 1.30 | 5.35 | No Bacterial pneumonia | moderate (3+) |
| Ji | 2018 | China | 359 | Positive | Mortality | Diagnosis delay>4 weeks | aHR | 2.60 | 1.42 | 4.78 | Diagnosis delay≤4 weeks | moderate (3+) |
| Ji | 2018 | China | 359 | Positive | Mortality | CD4 <50 cells/μL | aHR | 2.38 | 1.27 | 4.43 | CD4+ T cell count ≥ 50 cells/mm3 | moderate (3+) |
| Kantipong | 2012 | Thailand | 331 | Positive | Mortality | Meningeal TB | aOR | 3.27 | 1.37 | 7.82 | Pulmonary TB | moderate (3+) |
| Kantipong | 2012 | Thailand | 331 | Positive | Mortality | Casual laborer and farmer | aOR | 0.43 | 0.19 | 0.94 | Other occupations | moderate (3+) |
| Kazempour Dizaji | 2018 | Iran, Islamic Rep. | 2299 | Mixed | Mortality | Male gender | RR | 5.50 | 2.20 | 13.50 | Female gender | high (4+) |
| Kazempour Dizaji | 2018 | Iran, Islamic Rep. | 2299 | Mixed | Mortality | Comorbidities | RR | 4.90 | 2.00 | 11.60 | No Comorbidities | moderate (3+) |
| Kazempour Dizaji | 2018 | Iran, Islamic Rep. | 2299 | Mixed | Mortality | Tobacco | RR | 3.30 | 1.20 | 9.40 | No Smoker | moderate (3+) |
| Kebede | 2021 | Ethiopia | 465 | Mixed | Mortality | COPD | aOR | 4.60 | 1.30 | 16.70 | No COPD | moderate (3+) |
| Kebede | 2021 | Ethiopia | 465 | Mixed | Mortality | Heart failure | aOR | 3.80 | 1.40 | 10.40 | No Heart failure | moderate (3+) |
| Kebede | 2021 | Ethiopia | 465 | Mixed | Mortality | HIV positive | aOR | 2.90 | 1.70 | 5.00 | HIV negative | moderate (3+) |
| Kebede | 2021 | Ethiopia | 465 | Mixed | Mortality | Smear positive | aOR | 2.30 | 1.30 | 4.20 | Negative smear for PTB | moderate (3+) |
| Khosa | 2020 | Mozambique | 69 | Mixed | Sequelae | Never smoked males | RR | 0.15 | 0.02 | 0.89 | Ever smoked males | high (4+) |

| | | | | | | | | | | | | |
|---|---|---|---|---|---|---|---|---|---|---|---|---|
| Kim | 2012 | Korea, Rep. | 269 | Unknown | Mortality | High blood urine nitrogen | aOR | 3.23 | 1.23 | 8.49 | Normal blood urine nitrogen | moderate (3+) |
| Koegelenberg | 2014 | South Africa | 83 | Mixed | Mortality | Lobar consolidation | aOR | 0.05 | 0.01 | 0.43 | No Lobar consolidation | high (4+) |
| Koegelenberg | 2014 | South Africa | 83 | Mixed | Mortality | Cavitation | aOR | 0.19 | 0.04 | 0.95 | No Cavitation | high (4+) |
| Kosgei | 2020 | Kenya | 9026 | Positive | Mortality | BMI >25 kg/m² | aHR | 0.41 | 0.22 | 0.77 | BMI <15 | moderate (3+) |
| Kosgei | 2020 | Kenya | 9026 | Positive | Mortality | BMI 18.5–24.9 kg/m² | aHR | 0.48 | 0.38 | 0.59 | BMI <15 | moderate (3+) |
| Kraef | 2021 | International | 740 | Positive | Mortality | MDR-TB | aHR | 2.06 | 1.06 | 4.01 | No MDR-TB | moderate (3+) |
| Kraef | 2021 | International | 740 | Positive | Mortality | CD4 Unknown | aHR | 2.32 | 1.47 | 3.66 | CD4 ≥ 200/mm4 | moderate (3+) |
| Kubiak | 2018 | South Africa | 683 | Positive | Mortality | LAM grade 3, 4, or 5 | aHR | 2.39 | 1.14 | 4.98 | Urine LAM-negative | moderate (3+) |
| Kubiak | 2018 | South Africa | 683 | Positive | Mortality | LAM positive | aHR | 2.32 | 1.28 | 4.20 | Urine LAM-negative | moderate (3+) |
| Lalla | 2020 | South Africa | 78 | Mixed | Mortality | Septic shock | aOR | 5.21 | 1.97 | 13.75 | No Septic shock | high (4+) |
| Lalla | 2020 | South Africa | 78 | Mixed | Mortality | Renal Insufficiency | aOR | 3.48 | 1.28 | 9.33 | No Renal impairment | moderate (3+) |
| Lalla | 2020 | South Africa | 78 | Mixed | Mortality | Immunosuppression (HIV not included) | aOR | 2.13 | 1.48 | 2.56 | No other form of immunosuppression | moderate (3+) |
| Lin | 2015 | Taiwan | 2576 | Unknown | Mortality | Age ≥85 years | aOR | 3.40 | 2.56 | 4.53 | Age 65–74 years | moderate (3+) |
| Lin | 2015 | Taiwan | 2576 | Unknown | Mortality | Malignancy | aOR | 2.75 | 2.01 | 3.75 | No Malignancy | moderate (3+) |
| Lui | 2014 | China | 349 | Mixed | Mortality | Immunosuppression (HIV included) | aHR | 3.35 | 1.76 | 6.39 | No Immunocompromised conditions | moderate (3+) |

| | | | | | | | | | | | | |
|---|---|---|---|---|---|---|---|---|---|---|---|---|
| Lumu | 2021 | Uganda | 1111 | Positive | Mortality | Previous TB | aHR | 2.12 | 1.16 | 3.85 | No TB history | moderate (3+) |
| Macpherson | 2011 | Zimbabwe | 216 | Mixed | Mortality | Age 25–34 years | aHR | 2.45 | 1.03 | 5.82 | Age<25 years | moderate (3+) |
| Macpherson | 2011 | Zimbabwe | 216 | Mixed | Mortality | Recent weight loss | aHR | 2.53 | 1.25 | 5.12 | No Recent weight loss | moderate (3+) |
| Macpherson | 2011 | Zimbabwe | 216 | Mixed | Mortality | CD4 50–99 cells/μL | aHR | 6.31 | 2.52 | 15.83 | CD4 count≥201 | high (4+) |
| Macpherson | 2011 | Zimbabwe | 216 | Mixed | Mortality | CD4 Unknown | aHR | 6.10 | 2.63 | 14.12 | CD4 count≥202 | high (4+) |
| Marcy | 2014 | Cambodia | 661 | Positive | Mortality | Age ≥40 years | aHR | 2.83 | 1.49 | 5.36 | Age≤29years | moderate (3+) |
| Marcy | 2014 | Cambodia | 661 | Positive | Mortality | Anemia (Hb ≤7g/dL) | aHR | 2.48 | 1.31 | 4.72 | Hemoglobin>10 | moderate (3+) |
| Marcy | 2014 | Cambodia | 661 | Positive | Mortality | Disseminated TB | aHR | 2.47 | 1.53 | 4.00 | Pulmonary TB | moderate (3+) |
| Marcy | 2014 | Cambodia | 661 | Positive | Mortality | BMI ≤16 kg/m² | aHR | 2.32 | 1.21 | 4.44 | BMI>18.5 | moderate (3+) |
| Martin-Iguacel | 2021 | Denmark | 217 | Positive | Mortality | CD4 <100 cells/μL | RR | 2.57 | 1.34 | 4.94 | CD4≥200 | moderate (3+) |
| Martin-Iguacel | 2021 | Denmark | 217 | Positive | Mortality | Social burden | RR | 5.78 | 3.35 | 9.95 | No social burden | high (4+) |
| Martin-Iguacel | 2021 | Denmark | 217 | Positive | Mortality | Disseminated TB | RR | 3.32 | 1.11 | 9.94 | Extrapulmonary TB | moderate (3+) |
| Martin-Iguacel | 2021 | Denmark | 217 | Positive | Mortality | TB >3 months after HIV | RR | 4.34 | 2.17 | 8.66 | TB was diagnosed at up to 1 month before or within 3 months after HIV diagnosis | moderate (3+) |
| Massavirov | 2021 | Uzbekistan | 808 | Positive | Unsuccessful treatment | Diabetes Mellitus | aOR | 5.16 | 1.77 | 14.98 | No Diabetes | high (4+) |
| Massavirov | 2021 | Uzbekistan | 808 | Positive | Unsuccessful treatment | EPTB | aOR | 2.21 | 1.38 | 3.53 | Pulmonary TB | moderate (3+) |

| | | | | | | | | | | | | |
|---|---|---|---|---|---|---|---|---|---|---|---|---|
| Mbatchou Ngahane | 2016 | Cameroon | 269 | Mixed | Sequelae | Fibrotic pattern | aOR | 3.54 | 1.40 | 8.95 | No Fibrotic pattern | moderate (3+) |
| Millet | 2011 | Spain | 762 | Mixed | Mortality | HIV positive and Drug addiction | aHR | 7.70 | 4.50 | 13.30 | HIV Neg- Non-IDU | high (4+) |
| Millet | 2011 | Spain | 762 | Mixed | Mortality | Age 41-60 years | aHR | 3.50 | 2.10 | 5.70 | Age<41 years | moderate (3+) |
| Moolphate | 2011 | Thailand | 5626 | Mixed | Mortality | HIV positive | aOR | 3.24 | 2.46 | 4.26 | HIV negative | moderate (3+) |
| Moolphate | 2011 | Thailand | 5626 | Mixed | Mortality | Age 26–35 years | aOR | 2.57 | 1.26 | 5.24 | Age 16–25 years | moderate (3+) |
| Moolphate | 2011 | Thailand | 5626 | Mixed | Mortality | Age 36–45 years | aOR | 2.53 | 1.23 | 5.21 | Age 16–25 years | moderate (3+) |
| Moolphate | 2011 | Thailand | 5626 | Mixed | Mortality | Age 46–55 years | aOR | 4.60 | 2.22 | 9.54 | Age 16–25 years | moderate (3+) |
| Moolphate | 2011 | Thailand | 5626 | Mixed | Mortality | Age 56–65 years | aOR | 8.76 | 4.15 | 18.45 | Age 16–25 years | high (4+) |
| Moosazadeh | 2014 | Iran, Islamic Rep. | 964 | Mixed | Mortality | Age ≥55 years | aHR | 8.30 | 4.60 | 14.80 | Age 15-34 years | high (4+) |
| Moosazadeh | 2014 | Iran, Islamic Rep. | 964 | Mixed | Mortality | Renal Insufficiency | aHR | 4.10 | 2.10 | 8.10 | No Renal disease | moderate (3+) |
| Moosazadeh | 2014 | Iran, Islamic Rep. | 964 | Mixed | Mortality | Previous TB | aHR | 2.90 | 1.80 | 4.80 | No Previous TB treatment | moderate (3+) |
| Moreira | 2018 | Brazil | 473 | Positive | Mortality | Hyperglycemia (>126 mg/dl) | aHR | 3.72 | 2.17 | 6.38 | No hyperglycemia | moderate (3+) |
| Muyaya | 2018 | Botswana | 300 | Positive | Mortality | OI other than TB | aHR | 8.50 | 4.00 | 18.40 | No Oi other than TB | high (4+) |
| Muyaya | 2018 | Botswana | 300 | Positive | Mortality | Age ≥60 years | aHR | 4.80 | 1.80 | 13.00 | Age<35 years | moderate (3+) |
| Muyaya | 2018 | Botswana | 300 | Positive | Mortality | Anemia (Hb <10 g/dL) | aHR | 2.44 | 1.30 | 4.60 | Hemoglobin≥10 | moderate (3+) |

| | | | | | | | | | | | | |
|---|---|---|---|---|---|---|---|---|---|---|---|---|
| Nagai | 2017 | Japan | 345 | Negative | Mortality | Respiratory failure | aHR | 3.82 | 2.32 | 6.29 | No Respiratory failure | moderate (3+) |
| Nagai | 2017 | Japan | 345 | Negative | Mortality | Dehydration | aHR | 3.77 | 2.29 | 6.20 | No Dehydration | moderate (3+) |
| Nagai | 2017 | Japan | 345 | Negative | Mortality | Orientation disturbance | aHR | 2.65 | 1.62 | 4.34 | No Orientation disturbance | moderate (3+) |
| Nagai | 2017 | Japan | 345 | Negative | Mortality | Age: > 70 (men), > 75 (women) | aHR | 2.22 | 1.22 | 4.02 | Age: ≤70 (men), ≤ 75 (women) | moderate (3+) |
| Nahid | 2011 | United States | 565 | Mixed | Mortality | Smear positive | aHR | 3.07 | 1.44 | 6.56 | Smear negative | moderate (3+) |
| Nahid | 2011 | United States | 565 | Mixed | Mortality | HIV positive | aHR | 2.57 | 1.17 | 5.64 | HIV negative | moderate (3+) |
| Nguyen | 2018 | United States | 6755 | Mixed | Mortality | Meningeal TB | aOR | 8.22 | 3.69 | 18.32 | No Meningeal TB | high (4+) |
| Nguyen | 2018 | United States | 6755 | Mixed | Mortality | Chronic kidney failure | aOR | 5.66 | 2.78 | 11.54 | No Chronic kidney failure | high (4+) |
| Nguyen | 2018 | United States | 6755 | Mixed | Mortality | HIV status Unknown | aOR | 3.51 | 2.48 | 4.98 | HIV negative | moderate (3+) |
| Nguyen | 2018 | United States | 6755 | Mixed | Mortality | Age ≥65 years | aOR | 11.93 | 7.59 | 18.76 | Age 15–44 years | high (4+) |
| Nguyen | 2018 | United States | 6755 | Mixed | Mortality | HIV positive | aOR | 3.36 | 2.08 | 5.41 | HIV negative | moderate (3+) |
| Nguyen | 2018 | United States | 6755 | Mixed | Mortality | TB-specific abnormality on CXR | aOR | 2.84 | 1.49 | 5.40 | No TB-specific abnormality on CXR | moderate (3+) |

| | | | | | | | | | | | | |
|---|---|---|---|---|---|---|---|---|---|---|---|---|
| Nguyen | 2018 | United States | 6755 | Mixed | Mortality | Miliary TB | aOR | 2.38 | 1.27 | 4.48 | No Miliary TB | moderate (3+) |
| Nguyen | 2018 | United States | 6755 | Mixed | Mortality | Resident of long-term care facility | aOR | 2.36 | 1.22 | 4.59 | Not Resident of long-term care facility | moderate (3+) |
| Nguyen | 2018 | United States | 6755 | Mixed | Mortality | Age 45–64 years | aOR | 3.80 | 2.46 | 5.87 | Age 15–44 years | moderate (3+) |
| Nguyen | 2019 | United States | 1227 | Mixed | Mortality | Chronic kidney failure | aOR | 7.00 | 3.59 | 13.62 | No Chronic kidney failure | high (4+) |
| Nguyen | 2019 | United States | 1227 | Mixed | Mortality | Age ≥65 years | aOR | 4.85 | 3.00 | 7.85 | Age<65 years | moderate (3+) |
| Nguyen | 2019 | United States | 1227 | Mixed | Mortality | Miliary TB | aOR | 3.79 | 1.41 | 10.21 | No Miliary TB | moderate (3+) |
| Nguyen | 2019 | United States | 1227 | Mixed | Mortality | Homeless | aOR | 2.88 | 1.05 | 7.88 | Not Homeless | moderate (3+) |
| Osman | 2021 | South Africa | 2551058 | Mixed | Mortality | Age ≥65 years | aHR | 5.22 | 5.10 | 5.33 | Age 15–24 years | high (4+) |
| Osman | 2021 | South Africa | 2551058 | Mixed | Mortality | HIV Positive not on ART | aHR | 2.66 | 2.62 | 2.69 | HIV negative | moderate (3+) |
| Osman | 2021 | South Africa | 2551058 | Mixed | Mortality | Age 45–54 years | aHR | 2.11 | 2.07 | 2.15 | Age 15–24 years | moderate (3+) |
| Osman | 2021 | South Africa | 2551058 | Mixed | Mortality | HIV status unknown | aHR | 2.17 | 2.13 | 2.20 | HIV negative | moderate (3+) |
| Osman | 2021 | South Africa | 2551058 | Mixed | Mortality | Age 55–64 years | aHR | 3.15 | 3.09 | 3.22 | Age 15–24 years | moderate (3+) |
| Osman | 2015 | South Africa | 93133 | Mixed | Mortality | Age 15–24 years and CD4 < 50 cells/µL | RR | 10.29 | 7.66 | 13.80 | Age 15–24 years HIV negative | high (4+) |

| | | | | | | | | | | | | |
|---|---|---|---|---|---|---|---|---|---|---|---|---|
| Osman | 2015 | South Africa | 93133 | Mixed | Mortality | Age 15–24 years and CD4 201–350 cells/µL | RR | 2.70 | 1.84 | 3.96 | Age 15–24 years HIV negative | moderate (3+) |
| Osman | 2015 | South Africa | 93133 | Mixed | Mortality | Age 15–24 years and CD4 51–200 cells/µL | RR | 5.00 | 3.74 | 6.68 | Age 15–24 years HIV negative | moderate (3+) |
| Osman | 2015 | South Africa | 93133 | Mixed | Mortality | Age 25–34 years and CD4 < 50 cells/µL | RR | 6.56 | 5.40 | 7.95 | Age 25–34 HIV negative | high (4+) |
| Osman | 2015 | South Africa | 93133 | Mixed | Mortality | Age 25–34 years and CD4 201–350 cells/µL | RR | 2.18 | 1.73 | 2.74 | Age 25–34 HIV negative | moderate (3+) |
| Osman | 2015 | South Africa | 93133 | Mixed | Mortality | Age 25–34 years and CD4 51–200 cells/µL | RR | 3.69 | 3.05 | 4.47 | Age 25–34 HIV negative | moderate (3+) |
| Osman | 2015 | South Africa | 93133 | Mixed | Mortality | Age 35–44 years and CD4 < 50 cells/µL | RR | 4.18 | 3.49 | 5.00 | Age 35–44 years HIV negative | moderate (3+) |
| Osman | 2015 | South Africa | 93133 | Mixed | Mortality | Age 35–44 years and CD4 51–200 cells/µL | RR | 2.67 | 2.25 | 3.17 | Age 35–44 years HIV negative | moderate (3+) |
| Osman | 2015 | South Africa | 93133 | Mixed | Mortality | Age 45–54 years and CD4 < 50 cells/µL | RR | 3.30 | 2.75 | 3.96 | Age 45–54 years HIV negative | moderate (3+) |

| | | | | | | | | | | | | |
|---|---|---|---|---|---|---|---|---|---|---|---|---|
| Osman | 2015 | South Africa | 93133 | Mixed | Mortality | Age 55–64 years and CD4 < 50 cells/µL | RR | 2.46 | 1.88 | 3.22 | Age 55–64 years HIV negative | moderate (3+) |
| Osman | 2015 | South Africa | 93133 | Mixed | Mortality | CD4 <50 cells/µL | RR | 3.83 | 3.48 | 4.21 | HIV negative | moderate (3+) |
| Osman | 2015 | South Africa | 93133 | Mixed | Mortality | CD4 51–200 cells/µL | RR | 2.21 | 2.02 | 2.43 | HIV negative | moderate (3+) |
| Pecego | 2016 | Brazil | 44 | Positive | Mortality | CD4 <50 cells/µL | aHR | 4.38 | 1.64 | 12.74 | CD4≥50 | moderate (3+) |
| Pednekar | 2012 | India | 148173 | Unknown | Mortality | BMI 17 to 18.5 kg/m² | aHR | 4.54 | 2.14 | 9.63 | BMI 25.0 to <30.2 | moderate (3+) |
| Pepper | 2015 | South Africa | 16209 | Mixed | Mortality | Age ≥60 years | aHR | 7.22 | 5.11 | 10.19 | Age<25 years | high (4+) |
| Pepper | 2015 | South Africa | 16209 | Mixed | Mortality | Age 45–60 years | aHR | 3.20 | 2.40 | 4.27 | Age<25 years | moderate (3+) |
| Podlekareva | 2016 | International | 1406 | Positive | Mortality | MDR-TB | aHR | 3.32 | 2.05 | 5.37 | No Multidrug resistance | moderate (3+) |
| Ko | 2017 | Taiwan | 9831 | Mixed | Mortality | Age ≥75 years | aHR | 11.93 | 9.50 | 15.00 | Age<45 years old | high (4+) |
| Ko | 2017 | Taiwan | 9831 | Mixed | Mortality | Cancer | aHR | 3.01 | 2.61 | 3.47 | No Cancer | moderate (3+) |
| Ko | 2017 | Taiwan | 9831 | Mixed | Mortality | Heart failure | aHR | 2.84 | 2.53 | 3.18 | No Heart failure | moderate (3+) |
| Ko | 2017 | Taiwan | 9831 | Mixed | Mortality | Cerebral vascular disease | aHR | 2.71 | 2.45 | 3.00 | No Cerebral vascular disease | moderate (3+) |
| Ko | 2017 | Taiwan | 9831 | Mixed | Mortality | Hypertension | aHR | 2.58 | 2.32 | 2.86 | No Hypertension | moderate (3+) |
| Ko | 2017 | Taiwan | 9831 | Mixed | Mortality | Renal Insufficiency | aHR | 2.56 | 2.22 | 2.96 | No Chronic kidney disease | moderate (3+) |
| Ko | 2017 | Taiwan | 9831 | Mixed | Mortality | Age 45–54 years | aHR | 2.19 | 1.63 | 2.95 | Age<45 years old | moderate (3+) |

| | | | | | | | | | | | | |
|---|---|---|---|---|---|---|---|---|---|---|---|---|
| Ko | 2017 | Taiwan | 9831 | Mixed | Mortality | Ischemic heart disease | aHR | 2.12 | 1.92 | 2.34 | No Ischemic heart disease | moderate (3+) |
| Ko | 2017 | Taiwan | 9831 | Mixed | Mortality | Age 55–64 years | aHR | 3.31 | 2.53 | 4.35 | Age<45 years old | moderate (3+) |
| Ko | 2017 | Taiwan | 9831 | Mixed | Mortality | Age 65–74 years | aHR | 5.35 | 4.20 | 6.82 | Age<45 years old | high (4+) |
| Qi | 2020 | China | 485 | Positive | Mortality | ART not initiated | aOR | 11.00 | 6.40 | 18.90 | ART | high (4+) |
| Qi | 2020 | China | 485 | Positive | Mortality | Disseminated TB | aOR | 2.49 | 1.46 | 4.23 | No Disseminated TB | moderate (3+) |
| Rodrigo | 2016 | Spain | 5182 | Mixed | Mortality | HIV positive | aHR | 3.36 | 1.85 | 7.17 | HIV negative | moderate (3+) |
| Rodrigo | 2016 | Spain | 5182 | Mixed | Mortality | Retired | aHR | 2.40 | 1.11 | 5.14 | Employment status Active | moderate (3+) |
| Rodrigo | 2016 | Spain | 5182 | Mixed | Mortality | Smear positive | aHR | 2.32 | 1.04 | 4.76 | Culture negative | moderate (3+) |
| Rossetto | 2019 | Brazil | 2419 | Positive | Significant morbidity | MDR-TB | RR | 3.94 | 1.97 | 8.81 | Status at completion Cure | moderate (3+) |
| Rossetto | 2019 | Brazil | 2419 | Positive | Mortality | Years of education completed<7 | RR | 3.94 | 2.26 | 7.09 | Years of education completed ≥12 | moderate (3+) |
| Rossetto | 2019 | Brazil | 2419 | Positive | Significant morbidity | Years of education completed<7 | RR | 3.47 | 1.97 | 6.29 | Years of education completed ≥12 | moderate (3+) |
| Rossetto | 2019 | Brazil | 2419 | Positive | Mortality | University or higher education | RR | 2.84 | 1.61 | 5.16 | Years of education completed ≥13 | moderate (3+) |
| Rossetto | 2019 | Brazil | 2419 | Positive | Significant morbidity | University or higher education | RR | 2.56 | 1.44 | 4.69 | Years of education completed ≥13 | moderate (3+) |
| Saiki | 2019 | Japan | 279 | Negative | Mortality | Respiratory failure | aOR | 5.63 | 2.10 | 15.10 | No Respiratory failure | high (4+) |
| Saiki | 2019 | Japan | 279 | Negative | Mortality | Dementia | aOR | 3.20 | 1.15 | 8.88 | No Dementia | moderate (3+) |

| | | | | | | | | | | | | |
|---|---|---|---|---|---|---|---|---|---|---|---|---|
| Saiki | 2019 | Japan | 279 | Negative | Mortality | Decreased activity of daily living | aOR | 2.14 | 1.10 | 4.16 | Normal activity of daily living | moderate (3+) |
| Sanhueza-Sanzana | 2021 | Chile | 17512 | Positive | Mortality | CD4 ≥500 cells/μL | aHR | 0.07 | 0.01 | 0.56 | CD4< 351 | high (4+) |
| Sanhueza-Sanzana | 2021 | Chile | 17512 | Positive | Mortality | CD4 351-500 cells/μL | aHR | 0.32 | 0.11 | 0.89 | CD4< 350 | moderate (3+) |
| Sawadogo | 2015 | Burkina Faso | 200 (100 cases and 100 controls) | Mixed | Significant morbidity | Comorbidities | aOR | 5.74 | 1.69 | 19.44 | No Underlying condition | high (4+) |
| Sawadogo | 2015 | Burkina Faso | 200 (100 cases and 100 controls) | Mixed | Significant morbidity | Use of traditional medicine/herbs | aOR | 2.97 | 1.12 | 7.85 | No use of traditional medicine/herbs | moderate (3+) |
| Schmaltz | 2012 | Brazil | 227 | Positive | Mortality | ART initiated | aHR | 0.14 | 0.03 | 0.69 | No HAART | high (4+) |
| Sileshi | 2013 | Ethiopia | 422 | Positive | Mortality | No CPT | aHR | 3.03 | 1.58 | 5.79 | Cotrimoxazole prophylactic therapy | moderate (3+) |
| Sileshi | 2013 | Ethiopia | 422 | Positive | Mortality | EPTB | aHR | 2.39 | 1.23 | 4.66 | Smear negative PTB | moderate (3+) |
| Sileshi | 2013 | Ethiopia | 422 | Positive | Mortality | Functional status Ambulatory | aHR | 2.10 | 1.22 | 3.62 | Functional status Working | moderate (3+) |
| Sileshi | 2013 | Ethiopia | 422 | Positive | Mortality | CD4 <75 cells/μL | aHR | 4.83 | 1.98 | 11.78 | CD4 count≥250 | moderate (3+) |
| Sileshi | 2013 | Ethiopia | 422 | Positive | Mortality | CD4 150-250 cells/μL | aHR | 3.07 | 1.33 | 7.07 | CD4 count≥250 | moderate (3+) |

| | | | | | | | | | | | | |
|---|---|---|---|---|---|---|---|---|---|---|---|---|
| Sileshi | 2013 | Ethiopia | 422 | Positive | Mortality | CD4 75-150 cells/µL | aHR | 3.57 | 1.48 | 8.61 | CD4 count≥250 | moderate (3+) |
| Sileshi | 2013 | Ethiopia | 422 | Positive | Mortality | ART initiated | aHR | 0.35 | 0.19 | 0.64 | ART Not Started | moderate (3+) |
| Silva | 2018 | Brazil | 924 | Positive | Mortality | Age ≥50 years | aHR | 2.52 | 1.39 | 4.59 | Age<50 years | moderate (3+) |
| Silva | 2018 | Brazil | 924 | Positive | Mortality | ART not initiated | aHR | 2.91 | 1.71 | 4.93 | ART | moderate (3+) |
| Singla | 2021 | India | 250 | Mixed | Mortality | PaO2< 70 mmHg | aOR | 4.58 | 2.11 | 9.93 | PaO2≥70 mmHg | moderate (3+) |
| Singla | 2021 | India | 250 | Mixed | Mortality | SpO2< 90% | aOR | 4.25 | 4.01 | 6.17 | SpO2≥ 90% | moderate (3+) |
| Singla | 2021 | India | 250 | Mixed | Mortality | BMI <16 kg/m² | aOR | 3.80 | 1.12 | 12.88 | BMI ≥16 kg/m² | moderate (3+) |
| Singla | 2021 | India | 250 | Mixed | Mortality | Systolic blood pressure < 90 mmHg | aOR | 3.16 | 1.64 | 6.08 | Systolic blood pressure ≥90 mmHg | moderate (3+) |
| Singla | 2021 | India | 250 | Mixed | Mortality | Cavitation | aOR | 2.99 | 1.12 | 7.95 | No Cavity | moderate (3+) |
| Singla | 2021 | India | 250 | Mixed | Mortality | Respiratory Rate > 20 per minute | aOR | 3.29 | 1.55 | 6.95 | Respiratory Rate ≤ 20 per minute | moderate (3+) |
| Singla | 2021 | India | 250 | Mixed | Mortality | Serum sodium, <135 mEq/L | aOR | 2.60 | 1.28 | 5.28 | Serum sodium, ≥135 mEq/L | moderate (3+) |
| Singla | 2021 | India | 250 | Mixed | Mortality | Heart rate > 100 per minute | aOR | 2.27 | 1.12 | 2.59 | Heart rate ≤ 100 per minute | moderate (3+) |
| Talbot | 2012 | Tanzania | 212 | Positive | Mortality | ART initiated | RR | 2.99 | 1.42 | 6.32 | No ART | moderate (3+) |
| Thomas | 2021 | India | 212 | Mixed | Mortality | PTB | aOR | 2.83 | 1.15 | 6.95 | Extrapulmonary and disseminated tuberculosis | moderate (3+) |

| | | | | | | | | | | | | |
|---|---|---|---|---|---|---|---|---|---|---|---|---|
| Thomas | 2019 | India | 455 | Mixed | Unsuccessful treatment | Past Alcohol use and Audit ≥ 8 | RR | 4.67 | 2.17 | 10.02 | Never Alcohol use and Audit <8 | moderate (3+) |
| Thomas | 2019 | India | 455 | Mixed | Unsuccessful treatment | Current Alcohol use and Audit ≥8 | RR | 3.58 | 1.89 | 6.76 | Never Alcohol use and Audit <10 | moderate (3+) |
| van der Heijden | 2012 | United States | 609 | Mixed | Mortality | HIV positive | aOR | 8.08 | 3.83 | 17.06 | HIV negative | high (4+) |
| van't Hoog | 2012 | Kenya | 9381 | Mixed | Mortality | Age ≥65 years | aHR | 3.10 | 2.30 | 4.20 | Age 15–24 years | moderate (3+) |
| van't Hoog | 2012 | Kenya | 9381 | Mixed | Mortality | HIV Positive not on ART | aHR | 3.30 | 2.40 | 4.50 | HIV-negative | moderate (3+) |
| van't Hoog | 2012 | Kenya | 9381 | Mixed | Mortality | HIV status unknown | aHR | 2.80 | 2.10 | 3.70 | HIV-negative | moderate (3+) |
| van't Hoog | 2012 | Kenya | 9381 | Mixed | Mortality | HIV-positive, ART unknown | aHR | 2.80 | 2.10 | 3.60 | HIV-negative | moderate (3+) |
| Workneh | 2016 | Ethiopia | 1314 | Mixed | Mortality | Diabetes mellitus | aHR | 3.96 | 1.76 | 8.89 | No Diabetes mellitus | moderate (3+) |
| Workneh | 2016 | Ethiopia | 1314 | Mixed | Mortality | HIV positive | aHR | 2.59 | 1.21 | 5.59 | HIV negative | moderate (3+) |
| Wu | 2021 | China | 311 | Positive | Mortality | Previous TB | aHR | 3.53 | 1.82 | 6.85 | No History of TB | moderate (3+) |
| Wu | 2021 | China | 311 | Positive | Mortality | ART not initiated | aHR | 2.94 | 1.31 | 6.63 | ART | moderate (3+) |
| Wu | 2021 | China | 311 | Positive | Mortality | Smear positive | aHR | 2.84 | 1.61 | 4.99 | Smear negative | moderate (3+) |
| Wu | 2021 | China | 311 | Positive | Mortality | Tobacco | aHR | 2.76 | 1.54 | 4.94 | No Tobacco use | moderate (3+) |
| Wu | 2021 | China | 311 | Positive | Mortality | PTB + EPTB | aHR | 2.39 | 1.37 | 4.18 | No Extrapulmonary involvement | moderate (3+) |
| Wu | 2021 | China | 311 | Positive | Mortality | CD4 ≤50 cells/μL | aHR | 3.45 | 1.95 | 6.10 | CD4>50 | moderate (3+) |

| | | | | | | | | | | | | |
|---|---|---|---|---|---|---|---|---|---|---|---|---|
| Wu | 2014 | Taiwan | 35260 | Mixed | Mortality | Renal Insufficiency | aOR | 2.58 | 2.23 | 3.00 | No Chronic kidney disease | moderate (3+) |
| Wu | 2014 | Taiwan | 35260 | Mixed | Mortality | X-ray Unknown/not performed | aOR | 2.41 | 1.68 | 3.46 | X-ray Normal | moderate (3+) |
| Xue | 2020 | China | 199 | Positive | Significant morbidity | Age 45–64 years | aOR | 0.36 | 0.13 | 0.99 | Age<25 years | moderate (3+) |
| Yen | 2013 | Taiwan | 4438 | Mixed | Mortality | Age ≥65 years | aOR | 3.60 | 2.57 | 5.03 | Age 18–64 years | moderate (3+) |
| Yen | 2013 | Taiwan | 4438 | Mixed | Mortality | Malignancy | aOR | 3.16 | 2.21 | 4.54 | No Malignancy | moderate (3+) |
| Yen | 2013 | Taiwan | 4438 | Mixed | Mortality | Renal Insufficiency | aOR | 2.49 | 1.32 | 4.71 | No End-stage renal disease | moderate (3+) |
| Yen | 2017 | Taiwan | 5011 | Unknown | Mortality | Age ≥85 years | aOR | 5.94 | 4.18 | 8.44 | Age 65–74 years | high (4+) |
| Yen | 2017 | Taiwan | 5011 | Unknown | Mortality | Smear positive | aOR | 2.49 | 1.91 | 3.23 | Smear negative | moderate (3+) |
| Yen | 2017 | Taiwan | 5011 | Unknown | Mortality | Renal Insufficiency | aOR | 2.34 | 1.37 | 4.00 | No End-stage renal disease | moderate (3+) |
| Yen | 2017 | Taiwan | 5011 | Unknown | Mortality | Age 75–84 years | aOR | 2.01 | 1.41 | 2.87 | Age 65–74 years | moderate (3+) |
| Yen | 2017 | Taiwan | 2410 | Unknown | Mortality | Age ≥65 years | aHR | 2.60 | 1.56 | 4.32 | Age<65 years | moderate (3+) |
| Yen | 2017 | Taiwan | 2410 | Unknown | Mortality | Unemployment | aHR | 2.60 | 1.27 | 5.31 | Employment | moderate (3+) |
| Yen | 2017 | Taiwan | 2410 | Unknown | Mortality | Malignancy | aHR | 2.30 | 1.60 | 3.31 | No Malignancy | moderate (3+) |
| Yen | 2017 | Taiwan | 2410 | Unknown | Mortality | Married | aHR | 2.28 | 1.16 | 4.45 | Unmarried | moderate (3+) |
| Yen | 2017 | Taiwan | 2410 | Unknown | Mortality | Smear positive | aHR | 2.23 | 1.63 | 3.06 | Smear negative | moderate (3+) |
| You | 2021 | China | 335 | Unknown | Unsuccessful treatment | Cavitation | aOR | 6.05 | 2.82 | 12.98 | No cavitation | high (4+) |

| | | | | | | | | | | | | |
|---|---|---|---|---|---|---|---|---|---|---|---|---|
| You | 2021 | China | 335 | Unknown | Unsuccessful treatment | No Physical activity, exercise | aOR | 5.96 | 1.93 | 18.45 | Physical activity, exercise | high (4+) |
| You | 2021 | China | 335 | Unknown | Unsuccessful treatment | Smear and culture negative | aOR | 3.37 | 1.44 | 7.92 | Smear and culture negative | moderate (3+) |
| Zhang | 2021 | China | 1007 | Positive | Mortality | Unexplained infections or space-occupying lesions | aHR | 7.49 | 5.34 | 10.49 | No Unexplained infections or space-occupying lesions | high (4+) |
| Zhang | 2021 | China | 1007 | Positive | Mortality | Malignancy | aHR | 4.87 | 2.79 | 8.50 | No Malignancies | moderate (3+) |
| Zhang | 2021 | China | 1007 | Positive | Mortality | Severe pneumonia | aHR | 4.84 | 3.00 | 7.81 | No Severe pneumonia | moderate (3+) |
| Zhang | 2021 | China | 1007 | Positive | Mortality | Meningeal TB | aHR | 2.11 | 1.40 | 3.18 | No Tuberculous meningitis | moderate (3+) |
| Zhang | 2021 | China | 1007 | Positive | Mortality | Hypoalbuminemia | aHR | 2.06 | 1.45 | 2.94 | No Hypoalbuminemia | moderate (3+) |
